# KBG Syndrome: Prospective Videoconferencing and Use of AI-driven Facial Phenotyping in 25 New Patients

**DOI:** 10.1101/2021.11.18.21266480

**Authors:** Lily Guo, Jiyeon Park, Edward Yi, Elaine Marchi, Tzung-Chien Hsieh, Yana Kibalnyk, Yolanda Moreno-Sáez, Saskia Biskup, Oliver Puk, Carmela Beger, Anastassia Voronova, Peter M. Krawitz, Gholson J. Lyon

## Abstract

Genetic variants in the gene Ankyrin Repeat Domain 11 (*ANKRD11*) and deletions in 16q24.3 are known to cause KBG syndrome, a rare syndrome associated with craniofacial, intellectual, and neurobehavioral anomalies. We report 25 unpublished individuals from 22 families, all with molecularly confirmed diagnoses of KBG syndrome. Twenty-one individuals have de novo variants, three have inherited variants, and one is inherited from a parent exhibiting low-level mosaicism. Of the variants, 20 are truncating (frameshift or nonsense), and the remaining five individuals have missense variants (with one of these in three family members). One of the missense variants has been found in at least two other affected individuals. We created a novel protocol for collection and reporting of data, including prospectively interviewing these individuals and their families throughout eight countries via videoconferencing by a single clinician. Participants’ medical records, including imaging, were reviewed, and data was uploaded to the Human Disease Gene website using Human Phenotype Ontology (HPO) terms. Photos of the participants were submitted to GestaltMatcher and Face2Gene (FDNA Inc, USA) for facial analysis, and we found similar facial phenotypes among the participants. Within our cohort, common traits included short stature, macrodontia, anteverted nares, wide nasal bridge, wide nasal base, thick eyebrows, synophrys and hypertelorism. Seventy-two percent of participants had gastrointestinal complaints and 80% had hearing loss. Three participants were started on growth hormone with positive results. Behavioral issues and global developmental delays were found in most participants. Neurologic abnormalities including seizures and/or EEG abnormalities were also very common (44%), suggesting that early detection and seizure prophylaxis could be an important point of intervention. Twenty-four percent were diagnosed with attention deficit hyperactivity disorder (ADHD) and 28% were diagnosed with autism spectrum disorder (ASD). Additionally, we have identified minimally reported symptoms, including recurrent sinus infections (16%) and previously unreported migraines (20%). Based on the videoconferencing and these data, we provide a set of recommendations regarding diagnostic and treatment approaches for KBG syndrome.

## Introduction

KBG syndrome (OMIM 148050) was first described by Herrmann et. al. in 1975 [1] and is named after the surnames (K-B-G) of the first families reported with the syndrome. The original report described anomalies such as short stature, skeletal abnormalities, cognitive disability, and specific craniofacial dysmorphisms. Subsequent research has validated these original findings and has also expanded the list of commonly seen anomalies, which include seizures, behavioral disturbances, and gastrointestinal issues [2]. Genetic variants in the gene Ankyrin Repeat Domain 11 (*ANKRD11*) and deletions in 16q24.3 are known to cause KBG syndrome [3]. One of the authors (GJL) was introduced to KBG syndrome by one of the original discoverers of the syndrome (John Opitz), and then published a case report describing a 13-year-old boy with epilepsy, severe developmental delay, distinct facial features, and hand anomalies [4]. In that case, the proband had his first epileptic episode at 3 years of age. After this episode, he lost all speech, began exhibiting autistic behavior, and also started to have frequent generalized tonic–clonic seizures. Over time, tonic, atonic, mild clonic, complex partial, myoclonic, and gelastic seizures were reported in the proband. Other developmental skills, including throwing a ball, responding to his name, feeding himself with utensils, and self-care skills were lost by 4 years of age. The very serious nature of his epilepsy and its impact on his development was notable. In the present study, the same clinician (G.J.L.) met and interviewed twenty-five individuals (11 females, 14 males) from 22 families with KBG syndrome, to investigate further the role of epilepsy and other conditions in possibly affecting the trajectory of neurodevelopment in these individuals. In addition, it was asked whether the current state of facial recognition software can play a role in the diagnosis of this syndrome, so efforts were made to test the current status of two leading algorithms in the field [5, 6]. In an ideal world, a facial photograph could be combined with medical records and variant prioritization efforts, after exome or whole genome sequencing, to more accurately classify new missense and other variants in rare syndromes as likely pathogenic. In the end, we demonstrated how to use the PEDIA approach [7] as a standard diagnostic workflow to perform variant prioritization that integrates phenotypic features, facial images, and exome data.

## Methods

Twenty-five individuals (11 females, 14 males) from 22 families were interviewed via Zoom by a single clinician (G.J.L.) over a 4-month period from February 2021 to June 2021. All patients were molecularly diagnosed with KBG syndrome. The individuals reside in eight different countries (which are not being listed on this MedRxiv preprint to help avoid the use of identifying information). The individuals were either self-referred or recruited via a private Facebook group created by the KBG Foundation. This Facebook group consists of the largest ever population of patients with KBG syndrome, with over 500 participating individuals, although the exact number with genetically identified KBG syndrome is unknown. Health information and photo consent was obtained from each participant.

Genetic reports, medical records including imaging and pictures were collected from the families by email and compiled prior to the interviews. All variants were annotated to the NM_013275.5 transcript in GrCh37/hg19. The duration of the interviews were approximately one to two hours long and consisted of the physician (G.J.L.) asking structured questions and visually assessing for facial and limb phenotypic characteristics. Every anomaly reported by these participants were compiled as standardized Human Phenotype Ontology (HPO) terms onto the open-source Human Disease Genes website series (HDG) [8]. The HDG is a research initiative that started at the Department of Human Genetics of the Radboud University Medical Center in Nijmegen, Netherlands, with the goal of collecting and providing clinical consequences of novel variants in the human genome on an international scale. Microsoft Excel (version 16.49) was then used to internally organize and quantify frequency of features observed. HPO terms were used to maintain a standardized documentation of phenotypic abnormalities. The presence of a trait or phenotype was documented as such when it was explicitly stated in the interview or found in the individual’s medical records. The absence of a trait was documented as such if the interviewee stated they did not possess a certain phenotype, if it was not mentioned within the interview, and/or if it was not found in the medical records.

Facial photos of the research participants, provided by the participants and/or taken by the clinician during videoconferencing, were loaded into two facial recognition websites, Face2Gene (algorithm version 20.1.4) [5] and GestaltMatcher (algorithm version 1.0) [6]. These programs use deep learning algorithms to build syndrome or patient classifiers.

Face2Gene is powered by DeepGestalt, a state-of-the-art facial phenotyping framework that measures the similarity between a patient and a genetic disorder. De-identified photos are input into deep convolutional neural networks that were trained by images of those with various genetic syndromes. The neural network then provides similarity scores of the information contained in the facial morphology (Gestalt scores) [5]. Syndromes with higher similarities to the input image are ranked higher on the prioritized list, allowing for artificial intelligence driven diagnosis. Moreover, Face2Gene allows the clinician or the individual uploading the picture to select relevant phenotypic features (i.e., anteverted nares, prominent nasal bridge, generalized hypotonia, etc.) that the patient possesses. The clinical note or any summary of patient information can be uploaded and used by the program to extract phenotypic features, which is then used to derive the Feature score, an indicator of how well the clinical text seems to fit a specific diagnosis. The most current version of Face2Gene (October 2021, algorithm version 20.1.4) was used, which provides rankings based on Gestalt and Feature scores, ranked High, Medium, and Low (**Figure 1**). The top-30 ranked syndromes are based on a “combined score” derived from the Gestalt and Features scores. This “combined score” is based on an optimization of a test set proprietary to FDNA. The clinician can confirm the diagnosis as being either in a “differential diagnosis”, “clinically diagnosed” or “molecularly diagnosed”.

**Figure 1:**
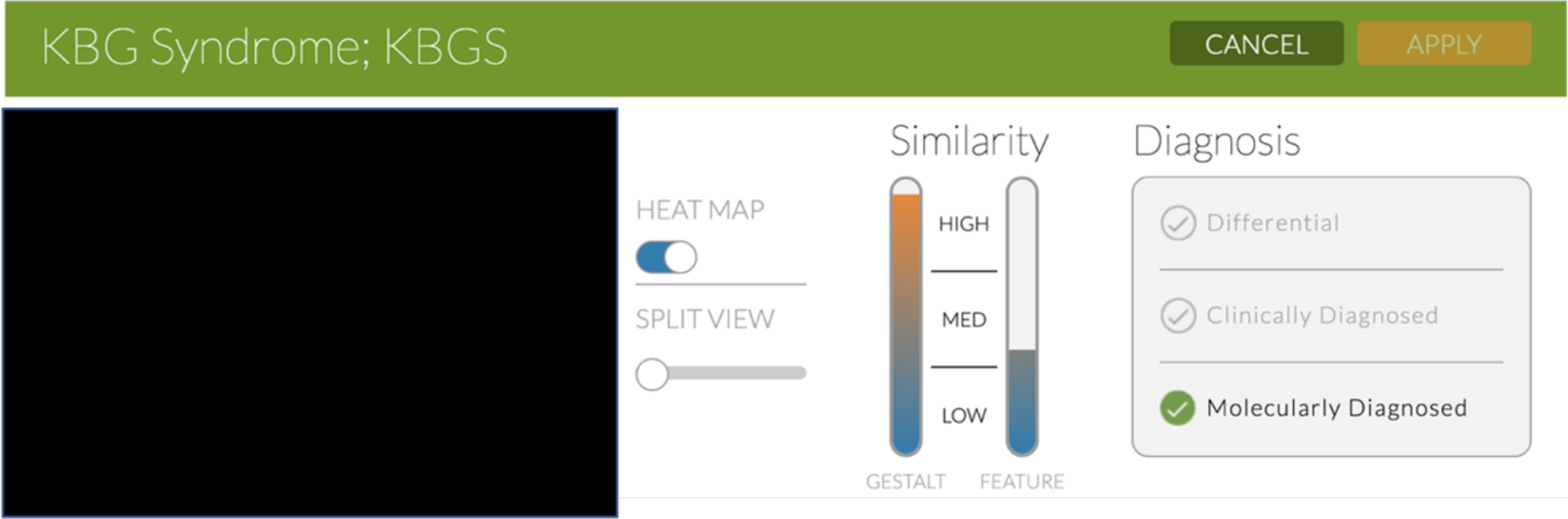
Face2Gene results of Individual R, indicating high Gestalt and medium Feature scores. Heatmap visualization shows the goodness-of-fit between areas of the individual image and each suggested syndrome. FOR MEDRXIV POSTING, THE PHOTOS HAVE BEEN REMOVED, AS THIS IS NOT ALLOWED BY THEM. CONTACT CORRESPONDING AUTHOR FOR THIS PHOTO.

GestaltMatcher, as an extension of DeepGestalt, can quantify the similarity between two patients, enabling one to identify genetic traits and associate them with the most likely genetic disorder. Our focus was on the clinical and phenotypic characteristics of KBG syndrome as presented in our cohort. GestaltMatcher spans a 320-dimensional clinical face phenotype space (CFPS) defined by the feature vectors derived from DeepGestalt [6]. Each image is a point located in CFPS, and the cosine distance quantified the similarity between two images in the space. In CFPS, we consider the images with close distance to have a high overlap of syndromic facial features. Therefore, we ranked the patients by sorting the cosine distance. We performed pairwise comparisons on the 25 photos of 25 participants (patients A-Y) against 3,533 images from 2,516 diagnosed patients with 816 syndromes in the Face2Gene database. These 2,516 patients were selected on the basis that their diagnosed syndrome was not included in the model training, and that there were less than seven participants with a given syndrome. By this selection criteria, we formed the CFPS with syndromes that the model has not seen and with very few participants simulating the ultra-rare diseases.

The main difference between Face2Gene and GestaltMatcher is that one quantifies the similarities on the syndrome level and the other on an image level. Given an image as input, Face2Gene quantifies the similarities to different disorders and returns a list of similar disorders. Conversely, GestaltMatcher quantifies the similarities between images and returns a score of similarity, given only one syndrome. In this case, we used GestaltMatcher to measure the similarities of the facial phenotypes between individuals with variants in *ANKRD11*.

The variant prioritization approach, PEDIA [7], takes a facial photo, clinical features (HPO terms) and exome sequencing data as input data for each patient. The current version of PEDIA (v1.1) integrates the score of each gene calculated from DeepGestalt, CADA [9] and CADD [10]. DeepGestalt first derives the gestalt score of each gene. The CADA score of each gene is calculated by the CADA web service (https://cada.gene-talk.de/webservice/) for the feature analysis. For the genomic score, since we currently only have the disease-causing mutation of each patient, we performed the exome simulation by inserting the disease-causing mutations into a randomly selected exome from the 1000 Genomes Project [11]. Then we annotated the exome variants with CADD score [10]. For the CADD score of each gene, we took the highest CADD score among the variants of each gene. We then trained the PEDIA model with the gestalt, CADA and CADD scores. For each patient, we obtained PEDIA scores for each gene. We then sorted the gene by PEDIA scores in descending order, and the top-1, top-10 and top-30 accuracy are reported in the results section.

## Results

We present 25 patients from 22 families with KBG syndrome, molecularly confirmed by identification of (likely) pathogenic variants in *ANKRD11.* We created a novel protocol that emphasizes data sharing, capitalizing on the use of video conferencing to reach a greater audience of those with rare genetic conditions, while helping train artificial intelligence software to aid in future diagnosis. The increased use and security of videoconferencing technology allowed access to participants in parts of the world outside of the United States, broadening the generalizability of our results. Given limited data and awareness for rare disorders, sharing data in a public domain such as the HDG website [8] which is accessible to families, researchers and health care providers, can further facilitate the development and refining of tools for health care providers to screen for rare disorders. A protocol that enables data sharing will also benefit AI software such as DeepGestalt, which requires thousands of patient images to model a vast range of genetic disorders and accurately train deep convolutional neural networks. Greater amounts of data and increased diversity of syndromes would improve the model’s generalization to rare disorders.

### Molecular Findings

Within our cohort of previously unpublished 25 KBG participants, twenty-one individuals have de novo variants, three have inherited variants, and one is inherited from a parent exhibiting a low-level of mosaicism in blood. Of these variants, 20 are truncating (frameshift or nonsense), and five are missense although one of these is in three members of one family. 21 distinct variants were identified. The location of the variants is shown in **Figure 2** and ages and the specific variants of the individuals and other characteristics, such as mode of inheritance, are shown in **Table 1** (**see excel sheet**). The truncating variants are classified by ACMG criteria [12] as: “PVS1 null variant (nonsense, frameshift) in a gene where LOF is a known mechanism of disease.” Furthermore, some variants are also classified as “PS2 De novo (both maternity and paternity confirmed) in a patient with the disease and no family history”. One of the missense variants p. (Val586Met) was observed in one heterozygous individual in Gnomad, and although the other two missense variants were absent from Gnomad, there were two heterozygous individuals in Gnomad with p. Arg2452His variant at the same amino acid position as p.Arg2452Gly observed here.

**Figure 2:**
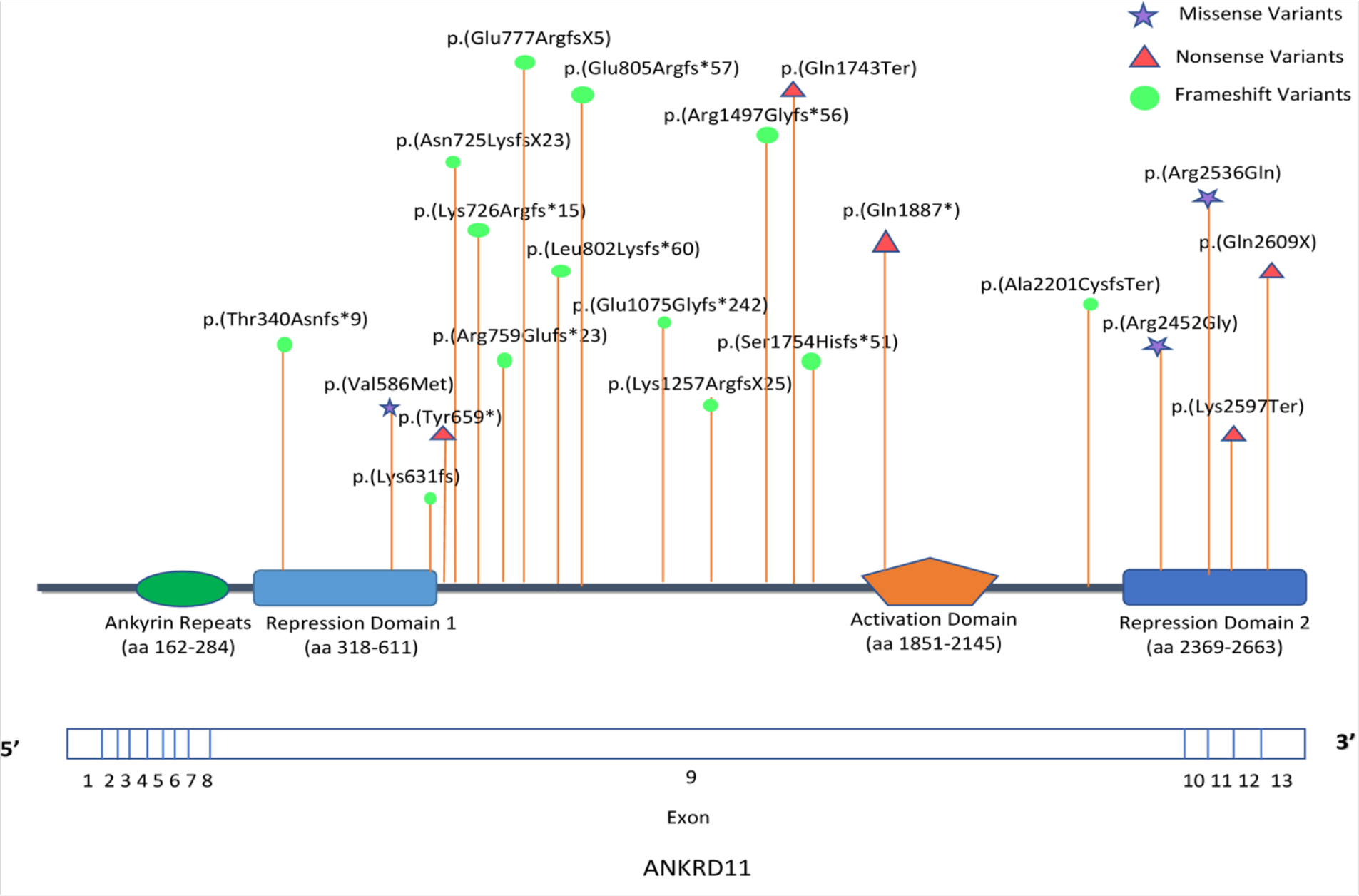
Schematic representation of DNA and protein variants of the 22 families along the *ANKRD11* gene. The coding exons for *ANKRD11* are depicted to scale. Abbreviations: aa = amino acid.

**Table 1:** See excel sheet. Cohort characteristics. This cohort includes 14 males and 11 females ranging from ages 1 year to 59 years. The mutation occurred de novo in 21 individuals and was maternally inherited in Individuals K and L, and paternally inherited in individual O. One parent of affected Individual T, Individual U, showed a low-level of mosaicism for the mutation.

Two of the missense variants (p.Arg2452Gly and p.Arg2536Gln) had been reported exactly once each in ClinVar, with p.Arg2452Gly reported by GeneDx (SCV000582346.5) and p.Arg2536Gln reported by CeGaT Praxis fuer Humangenetik Tuebingen (SCV001500097.3). In the case of Family C, with p.Arg2452Gly, this appears to be the same family already reported to ClinVar, as the sequencing was also performed by the company GeneDx, and GeneDx confirmed that they have only reported out one case with this variant on clinical exome sequencing, so this is the same case.

In the case of Family I, with the p.Arg2536Gln variant, this appears to be a new family, different from the one reported in ClinVar, as the sequencing was performed by the company Sistemas Genomicos, and Family I did not share their data with CeGaT Praxis fuer Humangenetik Tuebingen. This was confirmed by contacts at Sistemas Genomicos and CeGaT Praxis fuer Humangenetik Tuebingen. This variant was initially classified as a variant of uncertain significance as it had not been previously reported at the time of sequencing, but it is now being reclassified because of new information available: two additional patients carrying the exact same variant. The Arg2536Gln variant has been also recently reported in an unrelated patient affected with KBG syndrome, as a *de novo* event [13]. This patient has the following clinical features: poor weight gain, feeding problems and wide fontanel/delayed closure. Moreover, the variant in ClinVar is in a 14-year-old female of Greek origin (living in Germany) with suspected syndromic disease, mild intellectual disability, microcephaly, familial short stature and some dysmorphic features (thick eyebrows, synophrys, prominent nasal tip, macrodontia, thin upper lip vermillion). In this case, the variant was inherited from the mother who has short stature and mild learning disabilities. The maternal grandmother has short stature and a learning disability but has not been tested for the variant. Additional clinical information about this individual (referred to as Individual Z) is provided in Supplementary Information, although this family was not available for a videoconference. Additionally, a different nucleotide change affecting the same amino acid has been reported in a different patient with KBG syndrome [14] ( https://hdl.handle.net/2066/227290). Taking into account all this information, and given that more extensive cosegregation of the Greek family is not available, the variant is reclassified as likely pathogenic, according to ACMG criteria.

It is notable that as of November 13, 2021, there are 350 putative missense variants in ANKRD11 submitted to ClinVar, with many of these listed as variants of uncertain significance (**Supplementary Table 1**).

The frequency of phenotypic abnormalities in 25 KBG participants is presented in **Table 2**, and extensive clinical details for each case are provided in **Supplementary Information (which for the MedRxiv preprint will be available by contacting the corresponding author)**. The median age among the 25 individuals interviewed was 11-years-old, with the average age being 15-years-old (range = 1 to 59 years). Of the families, one reported coming from a consanguineous family, roughly half (n= 12) of the individuals had a history of congenital abnormalities within the family, and eight had a history of or relatives with intellectual disabilities. The parents of individuals B, D, T, and Y have had histories of miscarriage, although the variant was *de novo* for individual B, the inheritance pattern was unknown for individuals D and Y, and the parent (U) for individual T was mosaic with only 2 out of 298 sequencing reads from blood supporting this missense variant as mosaic in this parent. The mother of the 14-year-old female of Greek origin with the maternally inherited Arg2536Gln variant also has a history of several miscarriages early in pregnancy around 6 weeks of age.

**Table 2:** Frequency of phenotypic abnormalities in 25 KBG video-conferenced participants

| **Distinct Facial Features** |  |
| --- | --- |
| Eyes |  |
| Thick/ long eyebrows | 56% (14/25) |
| Synophrys | 44% (11/25) |
| Long/Prominent eyelashes | 32% (8/25) |
| Hypertelorism | 28% (7/25) |
| Strabismus | 28% (7/25) |
| Nose |  |
| Wide nasal bridge/ nose | 64% (16/25) |
| Prominent/broad nasal tip | 60% (15/25) |
| Anteverted nares | 52% (13/25) |
| Abnormal/wide nasal base | 32% (8/25) |
| Mouth |  |
| Macrodonia | 64% (16/25) |
| Thin upper lip vermillion | 32% (8/25) |
| Persistence of primary teeth | 16% (4/25) |
| Tented philtrum | 12% (3/25) |
| **Severity of Intellectual Disability** | **72% (18/25)** |
| None | 8% (2/25) |
| Mild | 56% (14/25) |
| Moderate | 12% (3/25) |
| Severe | 4% (1/25) |
| **Behavioral Abnormalities** | **84% (21/25)** |
| Abnormal mood | 48% (12/25) |
| Aggressive violent behavior | 28% (7/25) |
| Autism Spectrum Disorder | 28% (7/25) |
| Self-injurious behavior | 20% (5/25) |
| Impulsivity | 16% (4/25) |
| Repetitive compulsive behavior | 8% (2/25) |
| **Cardiac Abnormalities** | **52% (13/25)** |
| Patent foramen ovale | 12% (3/25) |
| Ventricular septal defect | 12% (3/25) |
| Persistent left superior vena cava | 8% (2/25) |
| Atrial septal defect | 8% (2/25) |
| **Neurological Abnormalities** | **96% (24/25)** |
| Developmental Delay | 80% (20/25) |
| Hypotonia | 56% (14/25) |
| Language Delay | 52% (13/25) |
| Seizures | 44% (11/25) |
| Gait disturbance | 40% (10/25) |
| Motor Delay | 32% (8/25) |
| Migraines | 24% (6/25) |
| ADHD | 24% (6/25) |
| **Skeletal Abnormalities** | **100% (25/25)** |
| Short Stature | 60% (15/25) |
| Clinodactyly | 48% (12/25) |
| Clinodactyly of the 5th digit | 50% (6/12) |
| Pes Planus | 40% (10/25) |
| Scoliosis (thoracic, lumbar, kyphoscoliosis) | 24% (6/25) |
| Sacral dimples | 20% (5/25) |
| Small hands | 16% (4/25) |
| Kyphosis | 16% (4/25) |
| Osteoporosis | 12% (3/25) |
| Abnormal or single palmar crease | 12% (3/25) |
| Abnormality of the ankle/Ankle clonus | 12% (3/25) |
| Abnormality of head shape | 16% (4/25) |
| **Gastrointestinal Abnormalities** | **80% (20/25)** |
| GERD | 44% (11/25) |
| Feeding difficulties in infancy | 44% (11/25) |
| Vomiting | 36% (9/25) |
| Chronic constipation | 28% (7/25) |
| NG/G tube feeding | 20% (5/25) |
| Abdominal pain/ migraines | 12% (3/25) |
| Hiatal/Inguinal hernia | 12% (3/25) |

### Cognition and Neurologic features

For the eight individuals who reported an intelligence quotient (IQ) score the mean IQ was 73 ± 4.84 (range= 64-80) as measured by the Weschler Intelligence Scale (3^rd –^ 5^th^ edition), except individual (T) who was administered the Reynolds Intellectual Assessment scales and received a composite intelligence score of 71. While the average composite IQ in the cohort is greater than what may be considered for an intellectual disability, factors such as overall functional status of the individual were considered. Overall, 68% of the cohort are considered mildly to moderately intellectually disabled based on their level of functioning. Global developmental delays prior to the age of 5 years were seen in 68% (n=17) of the group, with nine of the cases being classified as mild. Learning disabilities such as dyslexia, dysmetria, dysgraphia, and dyscalculia were observed in three separate individuals.

Eighty-eight percent of the participants (n=22) reported motor delays and 72% reported having speech delays (n=18). The known median age (reported by eight individuals) for when the child first started crawling was 12-months-old (range= 9-24 months). The known median age of onset of walking (reported by 10 individuals) was 22 months (range=12.5- 36 months), and the known median age of speech onset (reported by six individuals) was 30 months (range= 19-36 months). Individual B had selective mutism in the context of anxiety, whereas individuals J and W had absent speech entirely. Fine motor development delay (n=2) and poor fine motor coordination were present (n=3). Thirty-two percent (n=8) experienced general poor coordination.

Within this cohort, a vast majority experienced neurological abnormalities. The most common symptom was hypotonia (including two cases of neonatal hypotonia) followed by issues with gait. This included gait disturbance, gait ataxia, gait imbalance, shuffling gait, and broad-based gait. Seizures were common (44%) and included myoclonic, tonic-clonic, and absence seizures. No one specific type of seizure was seen. Electroencephalogram (EEG) abnormalities were documented in three of 11 individuals with seizures. According to maternal report, Individual E was meeting all motor and speech milestones (i.e., babbling, rolling over, and crawling) until the onset of myoclonic seizures, complex partial seizures, and verbal tonic seizures with respiratory distress around 0.5-2 years of age.

Similarly, individuals H, K, R, S, T, U, X, and Y reported histories of various types of seizures and concurrent speech and motor delays. Approximately a quarter reported migraines and one individual reported recurring headaches. Memory impairment was seen in two individuals, and dementia and brain atrophy were documented in one individual (P). Other brain abnormalities detected by magnetic resonance imaging (MRI) included pineal cyst, arachnoid cyst, choroid plexus cyst, subdural hemorrhage, and small pituitary gland.

Twenty-four percent (n=6) of individuals reported having a diagnosis of attention deficit hyperactivity disorder (ADHD), and two others were not given formal diagnoses of ADHD but reported having short attention span. Two individuals had concurrent diagnoses of cerebral palsy (8%) and seven had diagnoses of autism spectrum disorder (28%). Other neurological abnormalities seen included: syncope (n=2), apnea (n=4), and dysarthria (n=2). Behavioral abnormalities were prevalent (84%). The most common disorders were related to mood such as abnormal emotion or affect, depression, and/or anxiety.

Aggressive or violent behavior was the second most reported symptom, followed by self-injurious behavior including self-biting. Hand flapping was also documented (n=3). Of note, impaired pain sensation (n=3) and tactile sensation (n=2) was reported. Individual E and O report absence of pain threshold or not experiencing pain. Individual O has a history of a fractured foot and a dislocated kneecap. Bone scan showed normal density. Individual Q and R report a high pain threshold and decreased pain sensation, respectively. There was also impaired tactile sensation in two individuals (M,S).

### Stature

Heights, weight, and head circumference at birth and at the time of the videoconference were obtained for each individual and grouped based on whether or not they were below the 3rd percentile, between 3rd and 98th percentile, or greater than 98th percentile.

Individuals with known height at the time of video conference clustered into 3-98^th^ percentile (44%), below 3^rd^ percentile (24%) and above 98^th^ percentile (12%). Median height is 140.0 cm ± 29.4 cm. Known weights at time of videoconference clustered into 3- 98^th^ percentile (48%), below the 3^rd^ percentile (20%), and above 98^th^ percentile (4%).

Median weight is 27.8 kg ± 29.1 kg. Notably, of the three individuals who had heights above the 98^th^ percentile at time of videoconference, one had been put on growth hormone for approximately 2-4 years (Individual J) (**Table 3)**. Known birth length clustered into 3- 98^th^ percentile (44%), above 98^th^ percentile (8%), and below 3^rd^ percentile (8%). Median length was 49.0 cm ± 6.3 cm. Lastly, known birth weight mainly clustered between 3-98^th^ percentile (64%) with 16% below the 3^rd^ percentile and with no known individuals above the 98^th^ percentile. Median birth weight was 3 kg ± 0.7 kg.

**Table 3:** Height and weight percentiles for KBG participants at time of videoconference and at birth.

| Individual | Current Height (cm) | Height percentile | Current weight (kg) | Weight Percentile | Birth Length (cm) | Length Percentile | Birth Weight (kg) | Birth Weight Percentile |
| --- | --- | --- | --- | --- | --- | --- | --- | --- |
| Individual A | 163 | 3 (-1.89) | 68 | 45 (-0.13) | "normal" | unknown | 3 | 18 (-0.90) |
| Individual B | 118 | 73 (+0.61) | 23 | 73 (+0.61) | 49 | 42 (-0.20) | 3.27 | 38 (-0.30) |
| Individual C | 127 | 4 (-1.80) | 23.5 | 2 (-2.02) | unknown | 99 | unknown | unknown |
| Individual D | 150 | 2 (-2.04) | unknown | unknown | unknown | unknown | unknown | unknown |
| Individual E | 99 | 3 (-1.89) | 18.2 | 53 (+0.08) | 50.8 | 69 (+0.49) | 3.46 | 53 (+0.08) |
| Individual F | 140 | <1 (-3.58) | 32 | <1 (-3.80) | 31 | <1 (-8.28) | 0.773 | <1 (-5.05) |
| Individual G | 113.4 | <1 (-3.91) | 22.7 | 1 (-2.22) | unknown | unknown | 2 | 1 (-2.50) |
| Individual H | unknown | unknown | unknown | unknown | unknown | unknown | 3.3 | 34 (-0.42) |
| Individual I | 105 | > 99 (+3.47) | 16 | 93 (+1.49) | 46 | 6 (-1.56) | 2.73 | 9 (-1.33) |
| Individual J | unknown | 110 | unknown | 55 | 43.18 | <1 (-2.61) | 3.65 | 57 (+0.18) |
| Individual K | unknown | unknown | unknown | unknown | 56 | 99 (+2.23) | 3.27 | 32 (-0.47) |
| Individual L | unknown | 95 | 11.8 | 24 (-0.71) | unknown | unknown | unknown | unknown |
| Individual M | 177.8 | 99(+2.25 SD) | "normal" | unknown | unknown | unknown | unknown | unknown |
| Individual N | 79.4 | 8(-1.40 SD) | 9.4 | 1(-2.26 SD) | 50.8 | 60 (+0.24) | 3.23 | 30(-0.53 SD) |
| Individual O | 157.48 | <1(-2.65 SD) | 68.03 | 45(-0.12 SD) | 47 | 12 (-1.18) | 2.8 | 11 (-1.22) |
| Individual P | 157.48 | <1(-2.66 SD) | 58.97 | 11(-1.22 SD) | unknown | unknown | unknown | unknown |
| Individual Q | unknown | unknown | unknown | unknown | unknown | unknown | 3.26 | 38(-0.31 SD) |
| Individual R | 162.6 | 46(-0.10 SD) | 58 | 52(+0.04 SD) | 46.5 | 9 (-1.32 SD) | 2.4 | 3(-1.95 SD) |
| Individual S | 129.54 | 2(-2.02 SD) | 37.6 | 52(+0.04 SD) | unknown | unknown | 3.03 | 23(-0.75 SD) |
| Individual T | unknown | unknown | unknown | unknown | 49.5 | 40(-0.25 SD) | 3.4 | 40(-0.26 SD) |
| Individual U | unknown | unknown | unknown | unknown | unknown | unknown | 1.9 | <1 |
| Individual V | unknown | 40-50 | unknown | 40-50 | 49.53 | 51(+0.03 SD) | 2.948 | 18(-0.91 SD) |
| Individual W | 117 | 5(-1.65 SD) | 18.9 | 2(-2.10 SD) | unknown | 9 | 3 | 18(-0.90 SD) |
| Individual X | 160.02 | 34 (-0.40) | 66.22 | 76 (+0.70) | 49 | 7 (-1.50) | 2.18 | 1 (-2.37) |
| Individual Y | 182 | 92(+1.43 SD) | 115 | >99(+3.80 SD) | 46.99 | 12 (-1.19) | 2.72 | 9 (-1.35) |

### Facial Features

Facial features are demonstrated in **Figure 3**. At least one distinctive facial feature common to KBG patients was present in every individual interviewed, except perhaps for the parent (M) of individuals K and L, with the missense variant p. (Val586Met). Some defining facial characteristics observed include thick eyebrows with synophrys, prominent eyelashes, a wide nose, thin upper lip vermillion, and macrodontia. Several participants had a triangular face or pointed chin (n=7) and a broad or prominent forehead (n=3).

**Figure 3:** Clinical features of 25 KBG individuals. Characteristic features include bushy eyebrows (a,c,d,e,i,k,m,o,p,r,t,u,v,y), long eyelashes (c,d,i,l,o,p,s,x,), triangular face (a,g,k,r,v) and most had a wide nasal bridge or tip and a thin upper vermillion. MEDRXIV DOES NOT ALLOW POSTING OF PHOTOS. INSTEAD, THESE PHOTOS HAVE BEEN UPLOADED TO GESTALTMATCHER, AT LINK: https://db.gestaltmatcher.org/ AND INDIVIDUAL LINKS ARE IN TABLE 1. THIS FIGURE WILL APPEAR IN THE FINAL PEER REVIEWED PAPER AS WELL.

### GestaltMatcher Results for Facial Features

The pairwise ranks of the 25 photos in **Figure 4** showed similar facial dysmorphism among most patients. In the gallery of 3,533 images with 816 different disorders and 25 KBG patients, fifteen out of 25 KBG patients had at least one other KBG patient in their top-10 rank, and 21 out of 25 patients had at least one patient in their top-30 rank. Moreover, other than U being an outlier, there was a huge cluster containing the set of patients with three sub-clusters (P, J, F, and M), (O, H, R, Y, V, G, and I), and (Q, S, D, and E). The reason why patient U was an outlier was probably because of the low-level mosaicism for this variant. However, no clear groups were seen when analyzed to see whether patient phenotypes cluster by the type of genetic variant (missense, frameshift, nonsense). In summary, the results suggested that most of the patients described in this analysis share a similar facial phenotype. With the GestaltMatcher approach and the facial photos of patients, we can quantify the similarities among the patients. We can further utilize the similarities for diagnosing the patient and analyze the shared facial phenotypes associated with the genetic traits. GestaltMatcher has the potential to increase the accuracy of KBG diagnosis when used in conjunction with genetic testing.

**Figure 4:**
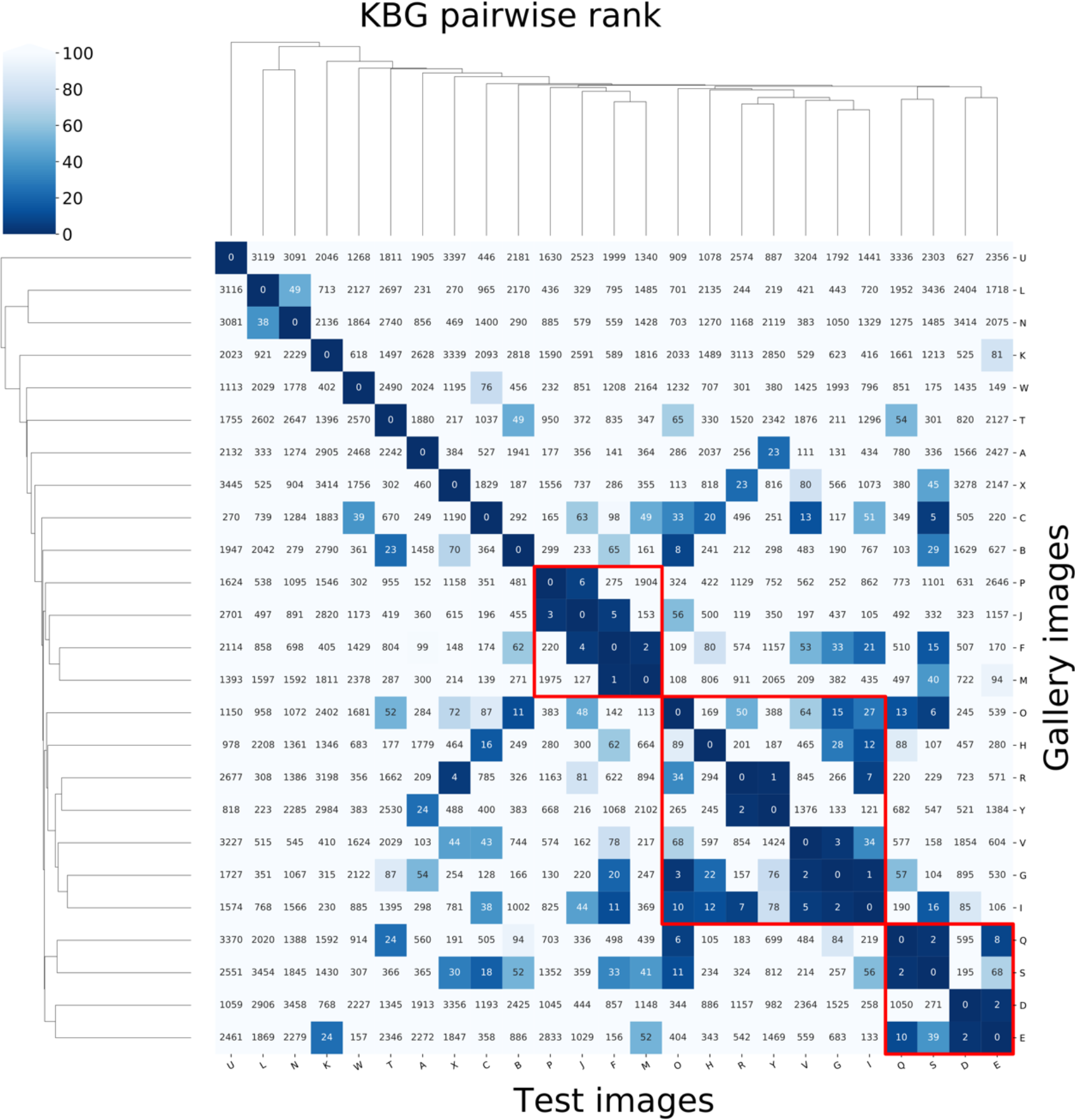
GestaltMatcher algorithm results; sub-cluster P, J, F, M all have synophrys or thick eyebrows, and a wide nose. Sub-cluster O, H, R, Y, V, G, I share common features of thick eyebrows, prominent or broad nasal tips, most had macrodontia, and several had a triangular face with a prominent forehead and pointed chin. Sub-cluster Q, S, D, E all have anteverted nares, broad nasal tip, and macrodontia. Individual E did not consent to having their photo published, however an actual frontal photo was input into the GestaltMatcher and Face2Gene algorithm.

Links to site: Individual A (https://db.gestaltmatcher.org/patients/5898), B (https://db.gestaltmatcher.org/patients/5899), C(https://db.gestaltmatcher.org/patients/5900), D (https://db.gestaltmatcher.org/patients/5901), E (https://db.gestaltmatcher.org/patients/5902), F (https://db.gestaltmatcher.org/patients/5903), G (https://db.gestaltmatcher.org/patients/5904), H (https://db.gestaltmatcher.org/patients/4683), I (https://db.gestaltmatcher.org/patients/5905), J (https://db.gestaltmatcher.org/patients/5906), K (https://db.gestaltmatcher.org/patients/5907), L (https://db.gestaltmatcher.org/patients/5908), M (https://db.gestaltmatcher.org/patients/5909), N (https://db.gestaltmatcher.org/patients/5910), O (https://db.gestaltmatcher.org/patients/5911), P (https://db.gestaltmatcher.org/patients/3992), Q (https://db.gestaltmatcher.org/patients/5912), R (https://db.gestaltmatcher.org/patients/5913), S (https://db.gestaltmatcher.org/patients/5914), T (https://db.gestaltmatcher.org/patients/5915), U (https://db.gestaltmatcher.org/patients/5916), V (https://db.gestaltmatcher.org/patients/5917), W (https://db.gestaltmatcher.org/patients/5918), X (https://db.gestaltmatcher.org/patients/5919), Y (https://db.gestaltmatcher.org/patients/5920)

### Face2Gene Results

Facial photos and phenotypic data of the 25 KBG individuals in this cohort was input into the Face2Gene database on August 24, 2021. KBG syndrome was ranked as the first and most likely diagnosis for 28% of individuals (n= 7). It was ranked second for 40% (n=10) and third or fourth for 12% (n=3). Overall, 80% (n=20) of patient’s photos analyzed had KBG syndrome ranked in their top five potential diagnoses. Among the 20 cases in which KBG was ranked in the top five, seven cases obtained the highest gestalt level whereas medium and low gestalt were obtained in ten and three cases, respectively. None of the cases ranked high for features of KBG, however 14 cases ranked medium, five ranked low, and one was unranked for features of KBG. The features and gestalt levels provided by Face2Gene are associated with the clinical text and the photos analyzed, respectively. Individuals B, F, and J initially submitted photos where they were wearing glasses. After running analyses on the faces without glasses, the ranking of KBG surprisingly dropped from 2 to 6 for individual B, from 2 to 3 for individual J, and it did not change for individual F. While KBG ranking fluctuated, the gestalt and feature levels did not change between the photos with and without glasses, for any of the three individuals.

Five individuals (K, L, M, P, U) did not have KBG syndrome appear as a differential diagnosis out of a list of 30 syndromes provided by Face2Gene. Their first ranked diagnoses included other developmental syndromes such as Cornelia de Lange, Williams Beuren, Rubenstein Taybi, Angelman, and mucopolysaccharidosis. Notably, Individual P was 55-60 years old at the time of the videoconference and first submitted a picture of himself around that age. When a new picture of him at a younger age was input into F2G, KBG syndrome emerged as the second-rank syndrome. Both Individual P and U (30-35 years old) have ages that fall above our median age of 11 years and the age at which most individuals are diagnosed with KBG syndrome. The DeepGestalt algorithm behind Face2Gene relies on the photos that it is trained on. If it was trained on data from a specific age group, it is likely that it is difficult for the software to extrapolate results. The other three individuals who were unranked (K, L, and M) are all from the same family, and possess the same missense mutation (**Table 1**).

### Ear, nose, throat (ENT) and vision

Phenotypic abnormalities of the eyes, ears and palate were documented. Vision impairment was common (n= 18). Seven individuals reported strabismus or monocular strabismus, seven reported myopia, and eight reported diagnoses of astigmatism. Thirty-two percent had low-set ears, prominent ears, posteriorly rotated ears, or an attached lobe as assessed on video. Bilateral conductive and/or sensorineural hearing impairment and low frequency hearing loss were seen in 12 individuals. Six had chronic otitis media, and only one of those six individuals did not have hearing impairment. Those experiencing chronic otitis media likewise had a preauricular pit, abnormal or blocked eustachian tubes, abnormality of the tympanic membrane, and increased size of nasopharyngeal adenoids. Individual B reported bilateral hearing loss until Eustachian tube insertion, and currently has normal hearing. Individual F was diagnosed with large vestibular aqueduct syndrome (with Mondini dysplasia), leading to sensorineural hearing loss of the left ear and reduced hearing in the right ear. CT of the paranasal sinuses also showed right choanal atresia. She received bilateral hearing aids at the age of 10-15 years and reports delayed receptive language skills. Individual V reported mixed sensorineural and conductive hearing loss bilaterally, worse in the right warranting a unilateral hearing aid. She reported structurally larger ear canals and had received bilateral myringotomies 2-3 times due to fluid accumulation. Speech was delayed at ∼2 years old and she has been diagnosed with expressive language disorder and receives special education and speech therapy while enrolled in a class for deaf children.

Within the study group, four individuals simultaneously experienced hearing loss and recurrent infections. Individual O was diagnosed with conductive hearing loss related to an abnormality of the eustachian tube and presence of a permanent hole in the eardrum of the contralateral ear. He has recurrent ear infections and has seen an immunologist due to spontaneous and recurrent fevers. Individuals P, Q, and Y have likewise been diagnosed with conductive hearing loss and reported recurrent sinus infections, chronic ear infections, and recurrent upper respiratory infections respectively. Lastly, palatal abnormalities such as high or narrow palate or soft and hard palate abnormalities were found in 24% (n=6) of the patients interviewed and of those six, four had difficulties feeding. Up to 50% of the group had other disorders of the face and jaw, such as ankyloglossia, retrognathia, and tongue thrusting. None of the families mentioned a small tongue, but this was not asked about specifically.

### Skeletal features

Skeletal abnormalities are a defining characteristic of KBG diagnosis, with the most common being short stature (60%). Clinodactyly and pes planus were particularly prevalent as well in 48% and 40% of participants, respectively. Abnormalities of head shape were observed including one individual with microcephaly and dolichocephaly, macrocephaly (n=2), and brachycephaly (n=1). Five individuals had delayed closure of the anterior fontanelles or abnormalities of the fontanelles. In regards to bone homeostasis, five within the cohort (Individuals A, F, H, T, Y) experienced reduced bone mineral density and have received diagnoses of osteopenia or osteoporosis. Individual A had an x-ray of his left hand and wrist which revealed physeal closure of the bones, excluding delayed bone maturation. A DEXA scan done in Individual F at 15-20 years revealed bone mineral densitometry below age range in the femoral head. Hand anomalies, mostly brachydactyly and/or clinodactyly of the 5th finger, have been previously reported in the majority of KBG syndrome patients [2]. Of the twenty-five interviewed, twelve had clinodactyly with half being of the 5th digit, and four had brachydactyly. One individual (S) reported a tethered spinal cord on MRI of the lumbar spine, and individual B reported spina bifida occulta. Twenty-eight percent (n=7) had sacral dimples while thirty-six percent (n=9) had scoliosis, kyphosis and/or lordosis.

### Cardiovascular features

Cardiac abnormalities were seen in approximately half the participants (**Table 2**). This included patent ductus arteriosus, patent foramen ovale, Tetralogy of Fallot, heart murmur, atrial fibrillation, and abnormal mitral valve morphology. Many of these abnormalities resolved over time, without the need for surgical intervention, although Individual K had Tetralogy of Fallot with pulmonary valve sparing surgical repair at ∼3-6 months of age, and Individual T had mitral valve repair at around one year of age.

### Gastrointestinal features

Gastrointestinal abnormalities were commonly experienced by 80% of the cohort. The most common issues were gastroesophageal reflux disorder (GERD) (44%) and feeding difficulties during infancy (44%). In addition, participants F, M, S, T, U had presumed diagnoses of abdominal migraines, characterized by stomach pain, nausea, and vomiting. Individual U reported abdominal pain and has been hospitalized for a history of migraines with aura and vomiting. She reports a weight loss of 41kg in the past year, and has other gastrointestinal symptoms such as gut spasms and constipation.

Individual Y reports history of migraines, episodic vomiting, as well as gastric immobility. He has been hospitalized 3-4 times for gastric impaction and constipation.

### Urogenital features

Other disorders such as urogenital disorders were seen in 48% (n=12) of individuals, with seven being female and five being male. Of note, four males were diagnosed with cryptorchidism. Other diagnoses included abnormalities of the urethra, recurrent urinary tract infection, bladder abnormalities, pollakiuria, polyuria, and enuresis. One individual was diagnosed with benign prostatic hyperplasia.

### Dermatologic features

A majority (56%) reported abnormalities of skin, nails, and hair, which included: hirsutism, low anterior hairline or abnormal hair whorl, cellulitis, keratosis pilaris, acne and dry skin, psoriasiform dermatitis, eczema, fingernail dysplasia, and recurrent fungal infections.

### Endocrinology, Metabolism, and Immune System function

Endocrine system disturbances were seen in 36% (n=9) and included abnormal glucose homeostasis, hyperglycemia and hypoglycemia, adrenal insufficiency, precocious puberty, elevated thyroid-stimulating hormone, and rheumatoid arthritis. Abnormalities of the metabolic system were seen in 64% (n=16), with slender build being the most common symptom (24%). Five reported a failure to thrive in infancy. One individual had blood disorders including iron deficiency anemia, reduced factor VII activity, and abnormal platelet function. Another individual reported abnormal circulating copper concentrations, and two others reported abnormalities in temperature regulation. Several experienced abnormalities of the immune system including allergies (n=11). One individual had abnormal immunoglobulin levels, immunodeficiency, pancytopenia, lymphopenia, abnormal B and T cell morphology as well as pneumonia, sinusitis, and esophagitis. Others suffered from recurring infections including that of the upper respiratory tract, asthma, and allergies including milk allergy.

Sixty-six percent of KBG individuals fall under the 10^th^ percentile for height [15]. This is consistent with our cohort where 60% reported short stature. Three individuals were administered growth hormone (Individuals H, J, O). Individual H was born at 3.3kg (34th percentile) at 38-40 weeks gestational age and he was diagnosed with a failure to thrive and feeding and swallowing difficulties in infancy. Individual J was born at 38-40 weeks with a weight of 3.65kg (57th percentile) and a birth length of 17 inches (<1st percentile). He was diagnosed with a failure to thrive at a check-up at 6-weeks of age and started growth hormone injections (Humatrope) at 3-5 years of age. He is weaning off of injections at 5-10 years old. His weight at time of videoconference is at the 55th percentile and his height is at the 110th percentile. His parents report growth hormone treatment to have been very successful. Patient O was given growth hormone starting from age 5-10 years until age 10-15 yeards with positive results as well. His birth weight was <1 percentile and current weight is within the 45th percentile (68.03kg), however his height still falls below the 1^st^ percentile (157.48cm).

### Variant prioritization with facial images

With PEDIA score, the disease-causing gene *ANKRD11* is ranked at the first place in 18 out of 25 cases (top-1 accuracy: 72%). When looking at the top-10 genes, *ANKRD11* is listed in the top-10 genes in 22 out of 25 cases (top-10 accuracy: 88%). All the cases have *ANKRD11* in their top-30 genes.

## Discussion

The main features of KBG syndrome consist of developmental delay, facial dysmorphism, large upper central incisors (macrodontia), and skeletal anomalies. It has been suggested that four out of eight major criteria, namely macrodontia, characteristic facial appearance, neurological involvement, delayed bone age, short stature, hand findings, costovertebral anomalies, and the presence of a first-degree relative with the syndrome, be present for the diagnosis [3]. Our findings are in line with that of previous literature and criteria set for KBG diagnosis (Table 2). Deletions of *ANKRD11* have previously been linked to specific facial dysmorphologies such as prominent forehead, arched eyebrows, large ears, and a pointed chin. Every individual in our cohort had at least one phenotypic characteristic consistent with KBG syndrome, however the disorder can be difficult to diagnose on facial phenotype alone. While some have a constellation of facial features typical of KBG syndrome, others may look very different. This is reinforced by the presence of three different clusters of similar facial characteristics within our cohort detected by DeepGestalt. There are also clear phenotypic overlaps with other genetic syndromes, most notably, Cornelia de Lange (CdLS) [16]. CdLS was listed as a differential diagnosis on Face2Gene for several of the KBG syndrome photos.

Given the rare nature of the disease with fewer than 200 cases cited in the literature to date, it is likely that KBG syndrome is misdiagnosed or underdiagnosed especially by those who do not encounter the disorder often. This, along with the mild and nonspecific features of the syndrome, make facial analysis software a helpful tool to aid in diagnosis.

Previous literature has reported the average ability of the system to identify a case of CdLS was 87%, compared with the experts’ average of 77%. Furthermore, when additional photographs were added to the system for increased machine learning, the detection rate of the system increased to 94% [17]. In addition to CdLS, cases of alcohol related neurodevelopmental disorder were identified by the system more efficiently than by manual methods [17]. Face2Gene has been used on an international scale in the United States, Canada, Japan and India, for example [18-21]. This is important to note since dysmorphologists have noted that some physical characteristics vary based on ethnicity, for example, the different Lip–Philtrum Guides for African American and white children [17]. Patient’s ethnicity may play a role in diagnostic accuracy, however performance of the algorithm in that regard is still unclear.

Within our cohort the algorithm for GestaltMatcher identified KBG syndrome as one of the top-10 most likely diagnoses in 60% of individuals, and correctly ranked KBG as top-30 for 84% of cases. Likewise, DeepGestalt correctly identified 80% of cases and ranked KBG as top-five in the list of potential diagnoses, consistent with the literature. It did “miss” five individuals with molecularly diagnosed KBG syndrome pointing to the need for greater data collection and more training. Additionally, the user-interface currently does not allow clinicians to parse out feature scores and gestalt scores, making it difficult to see how much weight is being put on facial features versus clinical phenotype. It might benefit clinicians if the scores were reported separately, along with the Combined Score (with an explanation for how that score is derived), or if these were reported in a more transparent way that allows users to understand how exactly rank and feature scores are being calculated. Nevertheless, facial recognition software can be a good adjunct for diagnosis and allow for more focused variant analysis [22]. Both DeepGestalt and GestaltMatcher have the potential to narrow down a vast range of possible diagnoses from lists of variants that might be generated from exome or whole genome sequencing. Moreover, we also demonstrated how we could use the PEDIA approach to diagnose these research participants. The PEDIA approach reported 72% of cases with *ANKRD11* correctly listed in the top-1 rank. It outperformed the approach only using facial photos with DeepGestalt, where only 28% of cases with KBG syndrome were listed in the top-1 rank, shown in the previous section (Face2Gene Results). Therefore, we envision that this approach, integrating facial, feature, and exome analysis, could be integrated into future diagnostic pipelines, but this will of course have to involve integration between the diagnostic genetic testing companies and the provision of facial photographs and detailed phenotypic features. Unfortunately, many current pipelines simply have the clinicians providing very few details about each case to the diagnostic testing company (such as simply only writing in “developmental delay” and nothing else, and with no photographs provided), so the workflows would have to be revised, with guidelines in place to protect patient privacy too.

*ANKRD11* has previously been implicated in craniofacial abnormalities and bone homeostasis in two different mouse models [23, 24]. Yoda mice heterozygous for the *ANKRD11* missense mutation (involving a G/C to A/T transition in exon 11 resulting in nonconservative amino acid change from glutamate to lysine) exhibited shortened snouts, wider skulls, and deformed nasal bones, underlined by altered morphology of frontonasal sutures and failure of interfrontal suture to close. Mice lacking *ANKRD11* specifically in the neural crest, which contributes to craniofacial development, displayed abnormal craniofacial gestalt, retrognathia, delayed fontanelle closure, tooth abnormalities and frequent cleft or submucosal cleft palate. This is similar to the craniofacial anomalies seen in KBG syndrome patients, particularly the failure of fontanelles to close, abnormal palate morphology correlated to difficulties in feeding, and abnormal craniofacial development. Of note, microcephaly, minimally cited in literature [25], was present in one individual (F) within our cohort.

It is speculated that the craniofacial abnormalities and inner ear malformations seen in KBG patients are tied to the high rates of hearing loss, particularly conductive hearing loss. This was observed in six individuals (24%), and sensorineural or mixed hearing loss was seen in three individuals (12%) consistent with previous literature [15, 26]. Imaging reports of the head ordered to assess proper bone growth of the ear were available from Individual F who was diagnosed with conductive and mixed hearing loss. CT of her paranasal sinuses showed right posterior choanal stenosis. It has also been speculated that hearing loss is associated with recurrent infections [15]. Craniofacial dysmorphisms may also affect the formation of the sinus canals leading to recurrent sinus infections. Individual H reported malformed sinus and ear canals with recurrent infections. Individual U has been diagnosed with chronic maxillary sinusitis and underwent repair of left sinus cavity and a nasal septum deviation surgery. CT scanning of the face revealed near complete opacification of the left maxillary sinus with small air-fluid level. However, 32% experienced conductive or sensorineural hearing loss without experiencing sinus infections and more evidence is necessary to establish correlation. Conversely, in the case of chronic otits media, 83% of those diagnosed with chronic otitis media also had hearing loss and there is a more likely correlation. Furthermore, given the role of *ANKRD11* with bone ossification, maturation, and remodeling, diagnoses of osteopenia and osteoporosis was also prevalent in this population (20%), and is unlikely to be age related. The median age of those diagnosed was 17 years (range= 9-22 years).

Gallagher et al. [27] demonstrated in the mouse model that *Ankrd11* is a nuclear coregulator in the brain that is essential for proper formation and location of neurons during cortex development. Consistent with its chromatin remodeling role, *Ankrd11* was found to bind regulatory elements of several key neurodevelopmental genes. Neural stem cells isolated from Yoda mice displayed aberrant histone acetylation and altered global gene expression. They were also able to identify ASD-like behaviors in Yoda mice, revealing a possible model for a subset of people with ASD [27]. Microdeletions at 16q24.3 encompassing *ANKRD11* and adjacent gene, Zinc Finger Protein 778 (*ZNF778*) have also been associated with autism spectrum disorder [28]. One group showed *Ankrd11* knockdown led to abnormal dendrite growth and development in mice and noted similar abnormalities when compared to mice with Rett, Down, and Fragile X syndromes [29].

A large exome sequencing study (35,584 total samples, of which 11,986 with autism spectrum disorder) found the rate of disruptive *de novo ANKRD11* variants to be moderate in autism spectrum disorder and high in neurodevelopmental disorders [30]. Moreover, Iossifov et al. report *ANKRD11 de novo* variants were frequent in their cohort comprised of 2,500 families, each having a child with an autistic spectrum disorder [31]. While none of the interviewed children appeared to have the severe form of autism as originally described by Leo Kanner [32], twenty-eight percent of our cohort had been given a diagnosis of autism spectrum disorder by previous providers. Notably, at the time of the interview, these individuals were interactive, social and maintained eye contact. This was somewhat surprising to one of us (GJL), as the first case he reported on with KBG syndrome did seem to have a form of severe autism, alongside the intellectual disability and epilepsy [4]. It seems warranted to conduct in the future quantitative studies, with rating scales, as part of a longitudinal natural history study, as a way to really elaborate what ASD symptoms manifest in KBG syndrome, independent of the intellectual disability. However, from an overall psychiatric perspective, eighty percent had behavioral abnormalities, including aggressive and violent behavior, impulsivity, or repetitive compulsive behavior, consistent with what has been previously reported [33].

We introduce a previously unrecognized feature of KBG syndrome, reduced pain sensation and impaired tactile sensation, and this is worth further investigation, starting with whether the reduced sensation is due to peripheral versus central neurological impairment. In addition to reduced or absent pain sensation, migraines and abdominal migraines, the latter of which is not a well-known phenomenon in itself, were both novel findings in 25% and 20% of our cohort, respectively. As part of the diagnostic criteria for abdominal migraines, anorexia, vomiting, headache, photophobia, or pallor (of which two must be present) must occur with at least five attacks of abdominal pain [34]. The pain is usually midline, periumbilical or poorly localized, and dull or sore in nature. One proposed theory for abdominal migraines is altered gut motility, which could be secondary to abnormal serotonin secretion in gastrointestinal neurons; abnormal serotonin secretion could also lead to visceral hypersensitivity [34]. The overall prevalence of abdominal migraines in childhood ranges from 2.4 to 4.1% and is more common in girls [34], but the syndrome can be under-recognized in the population. Lastly, the presence of tethered spinal cords has not been commonly cited previously, but can be related to gait abnormality and urinary symptoms (as was seen in individual S). Sacral dimples are commonly seen in our cohort (20%) and can potentially be a sign of a tethered spinal cord. Of note, 36% of our cohort had kyphosis, scoliosis, or lordosis.

Seizures can be seen in up to 50% of the KBG population, and this is so prevalent that it has been argued that diagnostic criteria for KBG syndrome should include neurological involvement (i.e., seizures, global developmental delay) [35]. Age of onset can range from infancy to the teens and the type of epilepsy is nonspecific, although tonic-clonic seizures are most commonly seen. We further speculate based on our data and review of the literature that age of onset of seizures may be inversely linked to severity of developmental delay. A previous review of 50 case reports revealed 13/46 with seizures and an average IQ of 58 [35], and the same paper reported two twin boys, the seizures first manifested in the first twin at age 8 years in the form of partial complex and absence seizures requiring management with antiepileptics and he continues to experience one to two absence seizures per month. IQ testing at 9 years of age showed an IQ of 50. His twin sibling likewise had his first seizure at 8 years of age and educational testing at 9 years revealed an IQ score of 55. Additional cases included one boy who experienced several episodes of seizures (associated with hypocalcemia) starting at 19 months and lasting until the age of 5 years. He had moderate psychomotor retardation, sat at 8 months, walked at 24 months, and spoke words at about 36 months [22]. Two individuals from a cohort of Italian patients suffered from transient pubertal seizures and grand mal seizures. The former had a mild cognitive disability but was socially poorly adapted, and the latter had a cognitive deficit with an IQ score of 75 [23]. In another case, two sisters both presented with generalized epilepsy with febrile seizures. The first had myoclonus-atonic type seizures with sudden falling of the head and trunk starting at 9 months old and was treated with sodium valproate (VPA) for approximately 4 years. Neuropsychological analysis at age 8 showed an IQ of 73. Her sister had her first febrile seizure at 12 months and was likewise treated with low-dose VPA for about 4 years. She presented with normal neuro-psychomotor and weight-height development, and an absence of dysmorphic and radiologic alterations [24].

A 6-yr-old Korean boy with features of KBG syndrome had partial seizures as indicated by the electroencephalogram but did not have a clinical seizure. He showed borderline intellectual disability and near-normal social function at 6 years with an IQ of 70, developmental delays in speech and motor functions, and he started walking at 20 months of age [25]. Another boy with diagnosed haploinsufficiency of ANKRD11 had grand mal seizures and was on medication until 5 years of age, after which he was seizure free. He was evaluated at the age of 9 and diagnosed with borderline intellectual disability (total IQ of 77 (WISC-R)). His motor development was delayed and he walked at the age of 2 years.

Non-specific EEG anomalies can occur in up to 72% of KBG patients and a quarter of them later develop seizures [21]. Prominent EEG changes, seizures and developmental delay in KBG syndrome can be explained by the important role of ANKRD11 as a transcriptional regulator analogous to the MeCP2 gene involved in Rett syndrome by interacting with histone modifying enzymes. It localizes to the nuclei of neurons and accumulates in discrete inclusions when neurons are depolarized (but has homogeneous diffuse distribution in the resting state) and plays a role in neural plasticity [26]. EEG patterns should be carefully checked because of the high likelihood of occurrence of seizures, especially if abnormal. Characterization of typical EEG abnormalities may also help to diagnose patients sooner. No known EEG pattern exists for KBG syndrome (unlike other childhood genetic disorders such as Rett Syndrome, Fragile X syndrome, Angelman syndrome), and readings have widely been described as described as nonspecific or of uncertain significance. Systematic and thorough descriptions and reporting of EEG abnormalities can guide the physician in diagnosis. Obtaining a baseline EEG is likely warranted, and although an ambulatory EEG for 24-72 hours is ideal, this may not be practical in all cases, so a routine EEG for 30 minutes to one hour could suffice. While further data is needed, the trajectory of those who have seizures compared to those who do not seems to differ- with those not experiencing seizures showing better outcomes. If seizures can be detected early and managed properly, developmental outcomes can perhaps be improved. Further analysis, likely in conjunction with a prospective natural history study, should look carefully at the presence and the age of onset of seizures and the future level of overall functioning. It also seems warranted to convene an international summit of KBG experts, in order to develop guidelines concerning recommendations for EEG monitoring and/or other more intensive monitoring related to seizure development, as there is currently no “consensus” or “guidelines” paper in the peer-reviewed literature about KBG syndrome, unlike the case with many other rare syndrome that have been known to medicine for a longer period of time, such as Tuberous Sclerosis [36] and Fragile X [37]. A summit of KBG experts could also address the types of therapy (speech, physical, occupational, etc.) and frequency that should be recommended.

Other variable features of KBG syndrome include congenital heart defects, cryptorchidism, kyphoscoliosis, and precocious puberty. KBG is associated with several types of heart conditions and oftentimes a diagnosis can be made from detection of newborn cardiac abnormalities. For example, individual B had a persistent left superior vena cava that was first detected at 20 weeks on fetal ultrasound which prompted karyotyping, but unfortunately no further genetic work-up until after birth. KBG diagnosis was eventually made by whole exome sequencing at five years of age This raises the question of whether antenatal ultrasound can play a safe and non-invasive role in the initial detection of KBG syndrome for the purpose of improving prenatal diagnosis and counseling. Overall, the extent and variety of reported deficiencies in the KBG syndrome patients could be attributed to the role of ANKRD11 as a chromatin regulator. Since ANKRD11 interacts with several key proteins of chromatin remodeling complexes, such as histone deacetylases and acetyltransferases, nuclear co-receptors, etc. and regulates global gene expression [27], it is not surprising that variants in *ANKRD11* affect development and/or function of multiple organs. It is unknown why different KBG syndrome patients tend to have variable number and severity of phenotypes and co-morbidities, although this is likely modified by different genetic backgrounds, different environments, and some level of stochasticity [38].

### Conclusion and Treatment Recommendations

- The frequency of intellectual disability and learning disabilities warrants early screening, identification, and intervention. Benefit has been found from occupational therapy, physical therapy, and special education classes. Based on our clinical data and literature review, early intervention in the form of physical therapy, occupational therapy, and speech therapy is recommended, when needed, and anecdotal reports from families indicate that a frequency of at least once-weekly is ideal, although this needs more formal study.
- In line with previous literature [39, 40], our results provide evidence for the potential utility of growth hormone treatment in KBG patients with short stature (under their target range). Based on the experiences of the families who have found benefit with growth hormone use for short stature and lean muscle mass with absence of adverse side effects, it is recommended that families with adolescents diagnosed with KBG syndrome consult with their physicians about the use of growth hormone. However, more systematic study is needed to develop guidelines regarding possible recommendations regarding the use of growth hormone.
- The high rates of neurological involvement, specifically seizures, in adolescents point at the possible utility of screening using EEG upon diagnosis and the importance of regular monitoring by a neurologist. Further research is warranted to justify mandatory EEG screening, but it is worth noting that other rare diseases with a high incidence of seizures do have formal recommendations for baseline EEG screening, such as with Tuberous Sclerosis [36].
- Given the prevalence of hearing loss and speech delays we recommend that children with *ANKRD11* variants undergo baseline auditory screening, along with starting speech therapy, if necessary.
- Most research is needed to investigate whether aggressive antibiotic treatment could prevent hearing loss in these patients, given that otitis media is a frequent finding in KBG patients.
- A lower threshold is warranted for the use of CT scanning to detect sinus abnormalities, since KBG syndrome is associated with craniofacial dysmorphism.
- Congenital cardiac anomalies are frequently seen, and patients may benefit from cardiac screening (including echocardiography) upon diagnosis.
- Research involving the etiology of GI related symptoms with KBG syndrome is minimal despite the high prevalence of such symptoms within our cohort (80%). Future clinical effort should focus on GI symptoms, including abdominal migraines.
- Given the often-mild cognitive deficits and subtle dysmorphic features, KBG syndrome can often be underdiagnosed. Artificial intelligence software has the potential to make diagnoses that may otherwise be missed. The combination of current data including mouse model data, AI data, and patient data and registries, can optimize the diagnosis of KBG syndrome and help develop further guidelines or treatment recommendations moving forward.

### Data Availability

Data generated and analyzed during this study can be found within the published article and its supplementary files. Additional data are available from the corresponding author on reasonable request. The exome sequencing data were generated as part of clinical testing, so the underlying raw data are not consented for deposition to a public database.

## Data Availability

The exome sequencing data were generated as part of clinical testing, so the underlying raw data are not consented for deposition to a public database

## Acknowledgements

We would like to thank all of the KBG syndrome families who participated in this study, along with Annette Maughan and the KBG Foundation for referrals. We also thank Karen Low, Charlotte Ockeloen, Elke de Boer, Tjitske Kleefstra and Nicole Fleischer for critical comments on the manuscript. The authors would like to thank the Genome Aggregation Database (gnomAD) and the groups that provided exome and genome variant data to this resource. A full list of contributing groups can be found at http://gnomad.broadinstitute.org/about.

## Author Contributions

GJL was responsible for all videoconferencing and primary data collection, with secondary summaries performed by EM, LG, JP, and EY. Data analysis was performed by LG, JP, EY and GJL. Facial recognition analyses were performed by TH, with input from PK. AV and YK provided input on the basic science implications. The first draft of the manuscript was written by LG, JP, and EY, with extensive editing thereafter by GJL, with input from all other authors.

## Ethical Approval

Both oral and written patient consent were obtained for research and publication, with approval of protocol #7659 for the Jervis Clinic by the New York State Psychiatric Institute - Columbia University Department of Psychiatry Institutional Review Board. Written family consent was given for publication of any photography of the children.

## Database Deposition and Access

The sequencing data were generated as part of clinical testing, so the underlying raw data are not consented for deposition to a public database.

