## Supplementary Information for "KBG Syndrome: Prospective Videoconferencing and Use of AI-driven Facial Phenotyping in 25 New Patients"

# A

| | |
|---|---|
| Gene Name: | ANKRD11 |
| NM_ | NM_001256182.1 |
| cDNA change: | c.1018dup |
| Protein change: | p.Thr340Asnfs*9 |
| Genomic position: | g.89351932dup |
| Hg build: | Unknown |
| Mode of inheritance: | Autosomal |
| Zygosity: | Heterozygous |
| Classification: | Pathogenic |
| Other Variants | None |
| Reason for referral | The proband is a 21-year-old male, initially evaluated for developmental and cognitive delays at 3 years of age. He carries a variant mutation p.Thr340Asn on the ANKRD11 gene. |

| Birth History and Vitals | |
|---|---|
| Pregnancy: | Mother engaged in appropriate prenatal care and no obstetric complications noted. |
| Birth (NVD or C-section): | NVD |
| Gestational Age | 40 weeks |
| Feeding: | dysphagia |
| Postnatal Period: | No neonatal issues at birth; dysmorphic appearance |

| *Facial abnormalities:* | |
|---|---|
| Ear: | None |
| Eye: | Large eyebrows |
| Nose: | Prominent nasal tip; broad nasal bridge; broad nasal base |
| Mouth: | Persistent primary teeth, macrodontia, long upper lip vermillion; tongue thrusting; |
| Other: | Pointed chin, broad forehead, triangular face |

| | |
|---|---|
| Developmental Behavior | ADHD, OCD like behavior with frequent hand washing, autism, hyperactive, easily loses temper, impulsive, short attention span, self-injurious behavior, tongue thrusting; repetitive behaviors; global developmental delays, severe, negative chromosomal analysis for Fragile X; stopped school in 9$^{th}$ grade and took a few courses in graphic design |
| Motor Delay | Fine motor delay |
| Speech Delay: | Yes, speech and language delay |
| IQ | IQ 75 |

| | |
|---|---|
| Intellectual Disability: | None, based on IQ, but adaptive functioning |
| Vision: | N/A |
| Hearing: | N/A |
| ENT: | N/A |
| Neurology: | Global developmental delay; motor delay; gait disturbance; balance disturbance; unilateral strabismus |
| Seizures/ Date of onset: | None |
| Neuroimaging: | MRI Brain normal |
| Cardiovascular: | EKG on 9/24/19 showed normal sinus rhythm, Echo showed ejection fraction of 64% with no regional wall motion abnormalities, trivial mitral regurgitation, mild tricuspid regurgitation, and pulmonary artery systolic pressure of 28 mmHg |
| Gastrointestinal: | Feeding difficulties; Moderate reflux esophagitis and small sliding hiatal hernia per endoscopy in 2019; constipation; |
| Respiratory: | None |
| Urogenital | N/A, renal ultrasound done on 3/26/18 was normal |
| Musculoskeletal: | Low total bone density meeting criteria for osteopenia, followed later by a diagnosis of osteoporosis, with bone mineral density showing T-scores of lumbar spine (-3.2), total hip (-1.4) and femoral neck (-1.9), thoracolumbar scoliosis and thoracic kyphosis confirmed on X-ray; small cervical ribs; vertebral body height loss T5 and T7; pain in bones; bilateral clinodactyly 5$^{th}$ finger; simian crease; broad forehead; short stature; brachycephaly; no broken bones |
| Skin/Hair/Nails: | Lesion on scalp, low hairline, frontal hair whorl |
| Endocrine: | Hirsutism |
| Immunological: | N/A |
| Allergies: | No known allergies |
| Metabolic: | N/A |
| Neoplasm: | None |
| Ob/Gyn: | N/A |
| Family History of congenital disorders: | Two maternal first cousins with dwarfism and two second cousins with microcephaly |
| Family History of intellectual disabilities | None |
| Consanguinity: | Yes |
| Medications: | Risperidone; Sertraline; Vitamin D supplementation; took one dose of Ritalin for ADHD but did not tolerate |
| Services | OT, ST, life skills starting at 3 years of age |
| Family History | Maternal bipolar disorder; Paternal grandmother has osteoporosis, sister has major depressive disorder with comorbid anxiety |

| Growth Parameters* | |
|---|---|
| Birth weight (percentile): | 3 kg. |
| Birth length: | Unknown |
| Birth head circumference: | Unknown |
| Current weight: | 68 kg. (<50%) |
| Current length: | 163 cm (<5%) |

| | |
|---|---|
| Current head circumference: | 55.2 cm |
| * Reference CDC Growth Charts-Simulconsult/measurement. N/A= not available | |

| |
|---|
| HPO terms not available on website: |

Standardized Testing:

- Verbal IQ 82
- Performance IQ 72

# B

| | |
|---|---|
| Gene Name: | ANKRD11 |
| NM_ | NM_031275.5 |
| cDNA change: | c.2329_2332del |
| Protein change: | p.Glu777ArgfsTer5, |
| Genomic position: | g. |
| Hg build: | GRCh37/hg19 |
| Mode of inheritance: | unknown |
| Zygosity: | heterozygous |
| Classification: | Pathogenic |
| Reason for referral | The proband is a 6-year-old Caucasian female with mild-moderate developmental delay and skeletal abnormalities. She carries a variant mutation p.Glu777ArgfsTer5 on the ANKRD11 gene. |

| Birth History and Vitals | |
|---|---|
| Pregnancy: | Increased monitoring due to mother's advanced age (40yr); mild hydrocephaly, around 20 weeks of pregnancy, that resolved itself. A persistent vena cava left superior was detected at 20 weeks by fetal ultrasound. |
| Birth (NVD or C-section): | emergency caesarian section |
| Gestational Age | Term, 40 weeks 1 day |
| Feeding: | feeding difficulties in infancy |
| Postnatal Period: | The birth complicated by the umbilical cord wrapped twice around the neck and green amniotic fluid. No obvious hypoxia; and APGAR score 9 and 10. |

| *Facial abnormalities: no dysmorphia* | |
|---|---|
| Ear: | none |
| Eye: | none |
| Nose: | Button nose; flat nasal bridge |
| Mouth: | Slightly tented philtrum |
| Other: | |

| | |
|---|---|
| Developmental Behavior | Mild developmental delays, poor coordination, balance coordination issues, with frequent falls. There are no reported issues with depth perception and the patient can walk up the stairs and takes the stairs down with two feet at a time. There is no obvious cortical visual impairment- it seems to be an issue with coordination. She can count to 20, knows her colors, and can speak. The mother suspects ADHD but the patient has not undergone formal testing: anxiety; anxious selective mutism; seems to be shy, is smiling a lot, and is very interactive. There is no obvious sign of autism. She makes good eye contact and is reported to be social in the nursery school with eight other children and |

| | |
|---|---|
| | has a close to a friend; can count to 20, knows her colors, much less extreme than other cases. |
| Motor Delay | Developmental milestones were delayed; sat at 13 months of age and walked at 22 months. The mother reports child's poor coordination, balance, with frequent falls. There are no reported issues with depth perception and the patient is able to walk up the stairs and takes the stairs down with two feet at a time |
| Speech Delay: | delayed speech. Started to speak aged 2 (also due to hearing loss between age 1 and 2 which was resolved with insertion of eustachian tubes) |
| IQ | IQ74 |
| Intellectual Disability: | Mild ID |
| Vision: | Myopia, astigmatism; wears eye plaster every day since she is 4 years old (initially for 2 hours, now 1 hour a day). |
| Hearing: | Bilateral hearing loss before Eustachian tube insertion; now normal |
| Neurology: | abnormal EEG reported starting at age 2. Denies history of epilepsy and seizures; EEG has been repeated every 6 months due to unusual activity with spike waves.<br>She has a sacral dimple. |
| Neuroimaging: | MRI at age 2 normal |
| Cardiovascular: | EKG normal; persistent vena cava left superior was detected at 20 weeks by fetal ultrasound. |
| Gastrointestinal: | Feeding difficulties in infancy; reports issues with feeding after birth that required a plaster cast to latch onto breast, supplemented with formula. picky eater, has trouble chewing; never required a feeding tube. The mother reports that she is just starting to develop her front teeth and she is slow to lose her baby teeth. The dentist has been consulted and is okay with the development. Suffers from constipation since she is 5 years old. She had feeding difficulties as baby and during infancy due to sucking, chewing and swallowing difficulties. |
| Respiratory: | |
| Urogenital | |
| Musculoskeletal: | mild scoliosis; spina bifida occulta; sacral dimple; pes planus and inward feet, and is not wearing serial casting, braces, or AFO. no issues with the stature predicted; hypotonia and floppy as infant; no stiffness and no issues with the calf muscles or Achilles tendon. She has small hands and short fingers. |
| Skin/Hair/Nails: | mild eczema (neurodermitis) and keratosis pilaris on cheeks, upper arms and thighs. low anterior hairline |
| Endocrine: | normal |
| Immunological: | normal |
| Allergies: | penicillin (Amoxicillin). |
| Metabolic: | none |
| Neoplasm: | no |
| Inheritance: | unknown |
| Family History of congenital disorders: | One prior chemical pregnancy, as detected by pregnancy test, with menstrual period after 6 weeks. |
| Family History of intellectual disabilities | no |
| Consanguinity: | no |

| | |
|---|---|
| Medications: | |
| Services | Occupational, physical therapy, and speech therapies |
| Family History | None with major intellectual disability. |

| **Survey & Parental replies to determine Eosinophil Esophagitis status:** | |
|---|---|
| Question: | Answer provided by parent: |
| 1)Any allergy diagnosed? – to what? | *1)None. Some lactose intolerance found on allergy testing but no complaints* |
| 2)Any acute skin reaction or swelling reaction to a food? | *2)no* |
| 3)Anaphylaxis | *3) None. Other than penicillin* |
| 4)Any episodes of diarrhea/vomiting or failure to thrive in infancy responsive to food exclusion | *4) no* |
| 5)Was there any failure to gain weight? | *5) no* |
| 6)Eczema – severity / extent | *6) Dry skin over last 2 years- gave cream; father also has it- may have gotten from dad (pilaris, chicken skin?) parents do not have eczema* |
| 7) Asthma – severity / preventer | *7)no* |
| 8) Allergic rhinitis | *8) no. Did have bronchitis once* |
| 9) Odynophagia | *9) no* |
| 10)Dysphagia; particularly meat and bread getting stuck. | *10) no* |
| 11) Is there a need to have food cut up very small? | *11) No issues. She cuts up own food for past 2 years* |

| Growth Parameters* | |
|---|---|
| Birth weight (percentile): | Wt: 3.27 kg. (16%) |
| Birth length: | Ht: 49 cm. (12%) |
| Birth head circumference: | OFC: 35cm. (25%) |
| Current weight: | Wt: 23 kg. ( 77%) |
| Current length: | Ht: 118 cm. ( 72%) |
| Current head circumference: | |
| * Reference CDC Growth Charts-Simulconsult/measurement. N/A= not available | |

Standardized Testing:
The child receives occupational therapy, physical therapy,and speech therapy.

C

| | |
|---|---|
| Gene Name: | ANKRD11 |
| NM_ | NM_013275.5 |
| cDNA change: | c.7354C>G |
| Protein change: | p.Arg2452Gly |
| Genomic position: | g. |
| Hg build: | GRC h37 |
| Mode of inheritance: | De novo |
| Zygosity: | heterozygous |
| Classification: | pathogenic |
| Reason for referral | Mild facial dysmorphia and moderate developmental delay; the proband carries a Arg2452Gly(CGC>GGC) variant on the ANKRD11 gene |

| Birth History and Vitals | |
|---|---|
| Pregnancy: | Regular prenatal care; |
| Birth (NSVD or C-section): | NSVD |
| Gestational Age | 40 weeks |
| Feeding: | unremarkable |
| Postnatal Period: | unremarkable |

| *Facial abnormalities:* | *Facial features reminiscent of Cornelia de Lange* |
|---|---|
| Ear: | preauricular pits, low set ears |
| Eye: | Full eyebrows; hypertelorism; long and prominent eyelashes; |
| Nose: | Prominent nasal tip; wide nasal bridge |
| Mouth: | Macrodontia; tented philtrum; prominent lower lip vermillion; |
| Other: | Small chin |

| | |
|---|---|
| Developmental Behavior | autism; ADHD; anxiety, defiance, hand flapping, avoids eye contact; low frustration level; impulsive; short attention span; intellectual disability moderate.; OCD; repetitive behaviors; Can be very uncooperative. Irritable, and gets angry, no temper tantrums, but irritable. Does not cooperate, autism diagnosis, but in mild range. Very focused on weather channel, special interests, would identify many cars starting at age 3 or 4. Very focused on cars. Was focused on many dates. Hyper-focused on garbage trucks and trains. Will watch train video obsessively, counting cars, for 15-20 minutes. Gives reassurance to him. |

| | |
|---|---|
| Motor Delay | Yes, floppy tone as baby; delayed milestones crawled at 11 months and walked at 22 months; poor fine motor (unable to tie shoelaces); poor coordination issues; Trouble swinging on a swing, delays with this. Not able to coordinate. Trouble throwing a ball. Hard time with complex movements. Poor coordination. Can write his name. Trouble tying his shoes, even at age 10. |
| Speech Delay: | Yes, mildly delayed speech; |
| IQ | IQ 70 |
| Intellectual Disability: | Yes, mild |
| Vision: | Horizontal nystagmus; astigmatism, denies impaired depth perception |
| Hearing: | normal |
| Neurology: | Global developmental delay, poor coordination; gait disturbance; Poor motor coordination; hypotonia; hypotonia in infancy; |
| Neuroimaging: | Brain MRI was normal, twice. First one at age 4, and then again at age 7years; Did contrast on second MRI but no DTI. |
| Cardiovascular: | EKG and ECHO normal |
| Gastrointestinal: | No swallowing issues as an infant.<br>Never any major choking; later GERD; vomiting episodes; |
| Respiratory: | Pneumonia, 2011 |
| Urogenital | normal |
| Musculoskeletal: | Sacral dimple, core hypotonia, hypotonia in infancy, upper body hypotonia, ankle clonus, clinodactyly; pes planus; mild short stature; ankle laxity |
| Skin/Hair/Nails: | Fair skin; |
| Endocrine: | elevated TSH |
| Immunological: | unremarkable |
| Allergies: | No known allergies |
| Metabolic: | Chronic fatigue; |
| Neoplasm: | no |
| Inheritance: | X-linked |
| Family History of congenital disorders: | Neither parent harbor the variant on the ANKRD11 gene; has muscular dystrophy in the family, with Limb Girdle 2A, in a cousin (sister's son). |
| Family History of intellectual disabilities | no |
| Consanguinity: | no |
| Medications: | Ritalin; Daytrana patch, at age 7; guanfacine, but suppressed appetite. |
| Services | Special Education; PT OT; ABA, vision therapist |
| Family History | Mother diagnosed as cystic fibrosis carrier; lives with parents & sister; mom’s sister’s son has LGMD 2A ie. Calpainopathy, third cousin on mom’s side had infantile spinal muscular atrophy, patient’s grandmother‘s cousin had some form of muscular dystrophy |

| Survey & Parental replies to determine Eosinophil Esophagitis status: | |
|---|---|
| Question: | Answer provided by parent: |
| 1)Any allergy diagnosed? – to what? | 1) not really, oral allergy to raw fruit |
| 2)Any acute skin reaction or swelling reaction to a food? | 2)no |

| | |
|---|---|
| 3)Anaphylaxis | 3) no |
| 4)Any episodes of diarrhea/vomiting or failure to thrive in infancy responsive to food exclusion | 4) parent's state seems low on growth curve. |
| 5)Was there any failure to gain weight? | 5) no |
| 6)Eczema – severity / extent | 6) no |
| 7) Asthma – severity / preventer | 7)no |
| 8) Allergic rhinitis | 8)no, does get congestion in allergy season |
| 9) Odynophagia | 9) no |
| 10)Dysphagia; particularly meat and bread getting stuck. | 10)no |
| 11) Is there a need to have food cut up very small? | 11)no |

| Growth Parameters* | |
|---|---|
| Birth weight (percentile): | Wt: |
| Birth length: | Ht: cm. (99%) |
| Birth head circumference: | OFC: not available |
| Current weight: | Wt: 23.5 kg. (26%) |
| Current length: | Ht: 127 cm. (27%) |
| Current head circumference: | OFC: 53 cm. (56%) |
| * Reference CDC Growth Charts-Simulconsult/measurement. N/A= not available | |

Standardized Testing: Not performed for this study

# D

| | |
|---|---|
| Gene Name: | ANKRD11 |
| NM_ | NM_ |
| cDNA change: | c.5233_5234del |
| Protein change: | p.Ser1745Hisfs*51 |
| Genomic position: | g. Unknown |
| Hg build: | GRCh37 |
| Mode of inheritance: | Autosomal dominant |
| Zygosity: | Heterozygous |
| Classification: | Pathogenic |
| Reason for referral | The proband is a 34-year-old female diagnosed with KBG syndrome, two years ago. She has had speech and language delays and an IQ of 80. |

| Birth History and Vitals | |
|---|---|
| Pregnancy: | No issues with pregnancy, no substance use |
| Birth (NVD or C-section): | N/A |
| Gestational Age | N/A |
| Feeding: | None |
| Postnatal Period: | N/A |

| *Facial abnormalities:* | |
|---|---|
| Ear: | None |
| Eye: | Synophrys, trichomegaly, thick eyebrows, slanting palpebral fissures |
| Nose: | Appears to have anteverted nares with broad nasal tip and broad nasal base |
| Mouth: | Macrodontia, thin upper lip vermilion. No palate deformities |
| Other: | None |

| | |
|---|---|
| Developmental Behavior | Has anxiety but social anxiety is worst aspect. Works as home care provider but cannot shop due to anxiety. Reports feeling depressed. Denies panic disorder. Can only work 10 hours per week and will get angry and overwhelmed with more. Would hit things out of anger. Unclear ADD/ADHD diagnosis. Supposedly needs to see a psychiatrist per neuropsychiatric testing for anxiety. Has associates degree in human services but dropped out before earning a bachelor’s degree. |
| Motor Delay | Started walking at 12-13 months, one month late for crawling |
| Speech Delay: | Speech therapy from 6 to 9, does currently speak with a lisp |
| IQ | IQ80 |
| Intellectual Disability: | None |
| Vision: | Myopia. No strabismus |

| | |
|---|---|
| Hearing: | None |
| Neurology: | Poor short-term memory, decreased concentration, headaches. |
| Seizures: | No seizures |
| Neuroimaging: | MRI brain showed atrophy in frontal lobes(?) |
| Cardiovascular: | Born with heart defect that resolved. No other issues |
| Gastrointestinal: | Appendectomy in 2010 and cholecystectomy in 2012 |
| Respiratory: | None |
| Urogenital | Overactive bladder |
| Musculoskeletal: | Clinodactyly of both 5$^{th}$ fingers and brachydactyly of all fingers, small feet, in-toeing on right corrected with brace, pelvic tilt (left higher than right), CT spine showed possible scoliosis. No sacral dimple, does not know if she has tethered cord |
| Skin/Hair/Nails: | Single transverse palmar crease on right |
| Endocrine: | None |
| Immunological: | None |
| Allergies: | May have food allergy but no formal diagnosis, allergic to sulfa drugs and penicillin (unknown reaction) |
| Metabolic: | None |
| Neoplasm: | None |
| Inheritance: | Autosomal dominant |
| Family History of congenital disorders: | Son has hypermobility |
| Family History of intellectual disabilities | May have intellectual disability on mother's side |
| Consanguinity: | N/A |
| Medications: | Oxybutynin, something for anxiety, was not given medication for ADHD as parents said no |
| Services | IEP for math and English after needing to retake 1$^{st}$ grade. Converted to all subjects when she got older, speech therapy |
| Family History | Mother has KBG, aunt and grandmother may have KBG but not tested. Mother had two stillborn babies. Mother may have anxiety disorder and memory loss. Son does well in school. Son also has emotional dysregulation. Father had coronary artery disease requiring stent placement, two paternal uncles had double bypass surgeries. |

| Survey & Parental replies to determine Eosinophil Esophagitis status: | |
|---|---|
| Question: | Answer provided by parent: |
| 1)Any allergy diagnosed? – to what? | 1) Sulfa drugs and penicillin |
| 2)Any acute skin reaction or swelling reaction to a food? | 2) Possibly but not diagnosed |
| 3)Anaphylaxis | 3) N/A |
| 4)Any episodes of diarrhea/vomiting or failure to thrive in infancy responsive to food exclusion | 4) N/A |
| 5)Was there any failure to gain weight? | 5) N/A |
| 6)Eczema – severity / extent | 6) No |
| 7) Asthma – severity / preventer | 7) No |
| 8) Allergic rhinitis | 8) No |
| 9) Odynophagia | 9) N/A |

| | |
|---|---|
| 10)Dysphagia; particularly meat and bread getting stuck. | 10) No |
| 11) Is there a need to have food cut up very small? | 11) N/A |

| Growth Parameters* | |
|---|---|
| Birth weight (percentile): | Wt: kg. (%) |
| Birth length: | Ht: cm. (%) |
| Birth head circumference: | OFC: cm. (%) |
| Current weight: | Wt: kg. ( %) |
| Current length: | Ht: 150 cm. (2%) |
| Current head circumference: | OFC: cm. ( %) |
| * Reference CDC Growth Charts-Simulconsult/measurement. N/A= not available | |

Standardized Testing:

Full scale IQ of 80
Verbal reasoning is low average; perceptual reasoning below average; memory tasks per WRAML2 are low average to below average; associative and integrative abilities are variable; processing speed is in the low average range; reading recognition is low average, math is average2

# E

| | |
|---|---|
| Gene Name: | ANKRD11 |
| NM_ | NM_031275.5 |
| cDNA change: | c.2404_2407del |
| Protein change: | p.Leu802Lysfs*60 |
| Genomic position: | g. |
| Hg build: | GRCh37/hg19 |
| Mode of inheritance: | De novo |
| Zygosity: | Heterozygous |
| Classification: | Pathogenic |
| Other Variants | - ABCC8; c.2995C>T; p.Arg99X; (hyperinsulinism); maternally inherited; **pathogenic**<br>- KCNQ3; c.2611C>G; p.Pro871Ala (benign familial neonatal seizures) autosomal dominant; heterozygous; maternal origin; uncertain significance<br>- Von Willebrand mutation; c.6158A>G; p.Asn2053Ser (Von Willebrand Disease); autosomal dominant/autosomal recessive; heterozygous maternal origin; uncertain significance |

| | |
|---|---|
| | - GATA1 c.433C>T; p.Leu145Phe; (X-linked recessive thrombocytopenia with/without dyserythopoietic anemia); denovo; uncertain significance |
| Reason for referral | ANKRD11 mutation |

| Birth History and Vitals | |
|---|---|
| Pregnancy: | Normal pregnancy |
| Birth (NVD or C-section): | NVD |
| Gestational Age | 41 weeks |
| Feeding: | feeding issues in infancy; not eating; put on NG tube for 3 months then G tube was placed; was given Lasix and enalapril |
| Postnatal Period: | Pericardial effusion at 7 weeks; pneumogram was normal with tachypnea; readmitted at 6-7 days old; denies hypotonia in infancy; normal APGAR scores 8&9 |

| *Facial abnormalities:* | |
|---|---|
| Ear: | N/A |
| Eye: | Thick eyebrows |
| Nose: | Upturned nose |
| Mouth: | Macrodontia |
| Other: | |

| | |
|---|---|
| Developmental Behavior | Friendly, outgoing, social and loves people; Mild global developmental delay related to amount of time in hospitals; No autism diagnosis; milestones were on-time, (rolling over, crawling, babbling) until seizures onset per mother report; not yet toilet trained |
| Motor Delay | Abnormal gait |
| Speech Delay: | Speech & language delay; 2-3 word sentences at 3 years- after VNS was able to speak full sentences and knows 100 words |
| IQ | Unknown |
| Intellectual Disability: | Undergoing cognitive testing currently; clear delay with school work |
| Vision: | None |
| Hearing: | None |
| ENT: | Reactive airway disease; esophageal manometry abnormal-weak swallowing apparatus; mild tracheomalacia; candida esophagitis, sinusitis; |
| Neurology: | 13 months, Myoclonic seizures, complex partial seizures, and verbal tonic seizures with respiratory distress. Refractory epilepsy; vagus nerve stimulator implanted at 3 years; syncope; sudden onset ataxia with poor coordination accompanied by muscle weakness; impaired selective motor control/hypotonia; central apnea; hypoxia; denies migraines; no pain threshold; sleep study normal |
| Seizures/ Date of onset: | 13 months (8/14/14) |
| Neuroimaging: | 2 MRI Brain reported normal; except for arachnoid cyst, small pituitary, possible cortical dysplasia |

| | |
|---|---|
| | First EEG normal, 2nd EEG 3 days later and subsequent EEGs were abnormal |
| Cardiovascular: | VSD as an infant that closed on its own; ASD; poor venous access; Broviac catheter placed on right chest- developed seroma and edema |
| Gastrointestinal: | Feeding issues; has G-tube; hernia repair surgery; gastroesophageal reflux; Nissen fundoplication surgery; aspiration with cyanosis; recently intestinal failure resulting in TPN/Lipids and IV meds; constipation; abdominal pain; vomiting |
| Respiratory: | Hypoxia; respiratory distress; asthma; no COVID exposure |
| Urogenital | Bladder problems - not toilet trained; leaky bladder; urinary tract infections |
| Musculoskeletal: | Hypotonia; denies tethered spinal cord; broke arm after fall, broke leg getting out of bed at 18 months, broke coccyx; delayed bone age, short stature; gait ataxia; grew 3cm in 3 months after starting TPN; bone density scan normal; |
| Skin/Hair/Nails: | Cellulitis; impetigo; cyst drainage from eyelid |
| Endocrine: | glucose instability (hyper/hypoglycemia - ranges from 40-300 mg/dL; hypothyroidism; adrenal insufficiency; possible pituitary gland hypoplasia; growth hormone levels normal |
| Immunological: | Immune deficiency; history of urosepsis; pneumonia; sinusitis; cytopenia; T-cell and B-cell dysfunction; insufficient Ig levels; on IVIG since age 2; NK cell lymphopenia; recurring Candida esophagitis infections; denies elevated lactic acid and elevated CRP; denies leukocytosis |
| Allergies: | None |
| Metabolic: | platelet dysfunction; macrocytosis; iron deficiency/anemia; spontaneous bleeding and bruising; Factor 7 deficiency; edema |
| Neoplasm: | None |
| Inheritance: | De Novo |
| Family History of congenital disorders: | N/A |
| Family History of intellectual disabilities | N/A |
| Consanguinity: | N/A |
| Medications: | Vimpat (seizures), Banzel (seizures), Hizentra infusions (immune deficiency), Singulair; hydrocortisone pump; Symbicort inhaler; daily fluconazole; Synthroid (hypothyroid), Amicar (bleeding) |
| Services | Physical therapy, speech therapy, occupational therapy since 1 year old |
| Family History | 4 older siblings;<br>- 3rd son has VSD and had open heart surgery- he developed SA node failure, pacemaker, cardiomegaly, he has not been tested or sequenced.<br>- Oldest daughter has mitral valve prolapse, VSD that closed by itself, syncope<br>- Mother has paroxysmal supraventricular tachycardia and no VSD or mitral valve prolapse according to echo done as adult; history of syncope- possible postural orthostatic tachycardia syndrome (POTS); hypoglycemia<br>- Paternal grandfather has syncope |

| Growth Parameters* | |
|---|---|
| Birth weight (percentile): | Wt: kg. (%) |
| Birth length: | Ht: cm. (%) |
| Birth head circumference: | OFC: cm. (%) |
| Current weight: | Wt:18.2 kg. (53 %) |
| Current length: | Ht: 99 cm. (3 %) |
| Current head circumference: | OFC: cm. ( %) |
| * Reference CDC Growth Charts-Simulconsult/measurement. N/A= not available | |

| HPO terms not available on website: |
|---|
| complex partial seizures<br>impaired selective motor control<br>intestinal failure resulting in TPN/Lipids and IV meds<br>impetigo |

Standardized Testing:

F

| | |
|---|---|
| Gene Name: | ANKRD11 |
| NM_ | NM_013275.5 |
| cDNA change: | c.2177_2178del |
| Protein change: | p.Lys0726Argfs*15 |
| Genomic position: | g.89350772_89350773del |
| Hg build: | GRch37 |
| Mode of inheritance: | De novo |
| Zygosity: | Heterozygous |
| Classification: | Pathogenic |
| Other Variants | None |
| Reason for referral | The proband is a 20-year-old female who is developmentally delayed and who harbors a Lys0726Argfs*15 variant on the ANKRD11 |

| Birth History and Vitals | |
|---|---|
| Pregnancy: | Born as set of quadruplets from IVF, decreased fetal movement noted; no hypoxia during delivery, polymorphic eruption, took Prednisone 5mg-30mg-40 mg; then Bethamethasone. Ventilin, to stop early contractions; was also on Valium 5 mg four times daily for contractions; Penicillin- for B Strep; Adalat 10 bid, to slow heart rate; baby aspirin; Fefol (folic acid & iron); Calcium, magnesium and zinc pre-natal supplements; no substance use but mother did have small amount of brandy |
| Birth (NVD or C-section): | C-section |
| Gestational Age | 29 weeks and 3 days |
| Feeding: | Vomiting with feeds, slow feeder as well, poor suck reflex |
| Postnatal Period: | Resided in NICU weeks 1-4, mild respiratory distress, anemia of prematurity requiring blood transfusion; the proband neonate aspirated and had ventilation support and was weaned to room air. Also had necrotizing enterocolitis. Had jaundice. |

| *Facial abnormalities:* | |
|---|---|
| Ear: | None |
| Eye: | Synophrys; hyperopia |
| Nose: | Right choanal atresia and deviated septum with surgical correction, anteverted nares, and wide nasal bridge; prominent nasal tip; broad nasal base |
| Mouth: | Macrodontia; wide spaced teeth; long upper lip vermillion; |
| Other: | None |

| | |
|---|---|
| Developmental Behavior | Global developmental delays, mild; anxiety in crowds, panic attacks, difficulty expressing feelings, difficulty with reading (minor), writing, and comprehension (biggest issue), good eye contact. Delayed language skills with receptive than expressive language; difficulty with content versus structure; difficulty with phonological awareness skills. Few episodes of hand flapping starting in 2017 with associated anxiety |
| Motor Delay | Significant motor delay. Delay with walking, did not crawl. Did not walk until 2.5 years. Also has fine motor skills delay. |
| Speech Delay: | None |
| IQ | IQ64 |
| Intellectual Disability: | Mild intellectual disability |
| Vision: | Wears glasses, Salzmann's nodular degeneration with nodule removal, may require corneal transplant. No issue with depth perception |
| Hearing: | Hearing aids at age 14 years, for large vestibular aqueduct syndrome (with Mondini dysplasia), sensorineural hearing loss in left, reduced hearing in right |
| ENT: | N/A |
| Neurology: | Dysarthria, migraine with aura, vestibular neuritis with vertigo, no hypotonia; four episodes of arm spasm/dystonia starting in 2016 |
| Seizures/ Date of onset: | No seizures but possible absence seizures |
| Neuroimaging: | MRI Brain, CT head |
| Cardiovascular: | Patent ductus arteriosus, no angiograms |
| Gastrointestinal: | Feeding difficulties in infancy; Crohn's disease, abdominal migraines associated with constipation requiring suppositories at the hospital, cyclic vomiting requiring hospitalizations, vomiting with feeds. Severe acid reflux, Barrett's esophagus. No hematemesis. No need for Heimlich. First endoscopy at age 2 or 3 years. Had Nissen fundoplication. Had abdominal ultrasound. Barium swallow on 3/4/02 showed GERD and discoordinate peristalsis. CT abdomen/pelvis on 1/12/12 showed hypoplastic left hepatic lobe, enlargement of right hepatic lobe with midline positioning of stomach and high positioning of the splenic flexure; superior mesenteric vein has small caliber and is anterior to superior mesenteric artery; findings are of unknown significance. |
| Respiratory: | Aspirated as neonate; intubated for respiratory support while in NICU |
| Urogenital | Right sided duplex renal collecting system. No urinary symptoms. |
| Musculoskeletal: | Abnormal gait, polyarticular arthritis with swan neck deformities, reduced core strength, overlapping toes, right $5^{th}$ digit clinodactyly, scoliosis, kyphosis, and sacralized L5, bilateral hindfoot valgus, bilateral pes planus hammer toes, bilateral medial malleolar hemi-epiphysiodesis with left triple arthrodesis; osteoporosis with low bone density ;arthritis since age 4; short stature, never treated with growth hormones; no sacral dimple; no pectus excavatum. |
| Skin/Hair/Nails: | Keratosis pilaris |
| Endocrine: | Osteoporosis, collarbone fracture |
| Immunological: | Juvenile Polyarticular Arthritis. |
| Allergies: | Lorazepam, diazepam, leflunomide, wheat, banana |
| Metabolic: | No mitochondrial diseases |
| Neoplasm: | None |

| | |
|---|---|
| Inheritance: | Unknown |
| Family History of congenital disorders: | None |
| Family History of intellectual disabilities | None |
| Consanguinity: | Unknown |
| Medications: | Esomeprazole, ranitidine, stemetil for vestibular neuritis, alendronate, adalimumab, folic acid supplements, Largactil, for cyclic vomiting, amitriptyline, CoQ10, super complex vitamin B, L-carnitine, ondansetron, Phenergan, Vitamin D and calcium, cortisone injection, flunarizine, lorazepam, valium, leflunomide, etanercept, methotrexate, Amoxil, Predmix, cyproheptadine, lansoprazole, sumatriptan, dexamethasone, cyclizine, oxycodone |
| Services | Special school education |
| Family History | The proband is one of quadruplets; Mother has scoliosis, migraines, atrial fibrillation, osteoarthritis, and psoriasis. No abdominal migraines in the family. She reside with her biological parents; one sister and two brothers, all of whom are normally developing. |

| Growth Parameters* | |
|---|---|
| Birth weight (percentile): | Wt: 0.773 kg (<1%) |
| Birth length: | Ht: 31 cm (<1%) |
| Birth head circumference: | OFC: 24.6 cm (<1%) |
| Current weight: | Wt: 32 kg (<1%) |
| Current length: | Ht: 140 cm (<1%) |
| Current head circumference: | OFC: 50 cm (<1%) |
| * Reference CDC Growth Charts-Simulconsult/measurement. N/A= not available | |

| HPO terms not available on website: |
|---|

Standardized Testing:

On WISC-IV: verbal comprehension index was borderline, perceptual reasoning was extremely low range, working memory was borderline, processing speed is average, and full scale IQ is borderline

# G

| | |
|---|---|
| Gene Name: | ANKRD11 |
| NM_ | NM_013278.5 |
| cDNA change: | c.4489_4490del |
| Protein change: | p.Arg1497Glyfs*56 |
| Genomic position: | g.89348460_8934861del |
| Hg build: | GRch37 |
| Mode of inheritance: | denovo |
| Zygosity: | heterozygous |
| Classification: | pathogenic |
| Reason for referral | The proband is a 10-year-old male with a history of premature birth, bilateral hearing loss, mild intellectual disability and developmental delay. He carries the ANKRD11 gene. |

| Birth History and Vitals | |
|---|---|
| Pregnancy: | Mother has epilepsy; on folic acid and Topiramate during pregnancy |
| Birth (NVD or C-section): | NVD, delivered in toilet |
| Gestational Age | 35 weeks (5 weeks premature) |
| Feeding: | Feeding difficulties in infancy |
| Postnatal Period: | Neonate had an infection at birth; Admitted to NICU for 2 weeks; fresh blood in aspirates; prolong jaundice in infancy received phototherapy |

| *Facial abnormalities:* | |
|---|---|
| Ear: | Low set ears |
| Eye: | Hypertelorism; myopia; slanting of the palpebral fissure; nystagmus of the left eye |
| Nose: | Wide nasal bridge; prominent nasal tip; |
| Mouth: | Bifid uvula; thin upper vermillion; His palate was soft, and the roof of mouth would collapse if take out uvula. Soft palate, macrodontia |
| Other: | Triangular face; mild facial dysmorphia |

| | |
|---|---|
| Developmental Behavior | Global developmental delays- walking and crawling; mild learning disabled; aggressive behavior; sensory seeking behaviors; short attention span; receptive attention skills; anxiety at school; generally a happy child; good expressive language skills; poor receptive speech; poor eye contact; requires a structured environment; speech and language delay; occasional echolalia and hand clapping and flapping (observed by teacher); limited range of facial expressions; the boy is toilet trained; Trouble with social |

| | |
|---|---|
| | interactions. Will not play with others. No issues with eye contact. Does self-stimulating behaviors. Obsessed with Rolls-Royces automobiles. Seems fascinated by certain topics. He is verbal and has many vocabulary words.<br>Seems to have slurred speech. Trouble articulating.<br>Did get testing for autism and said he does not have autism. Has not yet had ADOS testing; has massive tantrums and hits himself on the head 15-20 times daily; quick to lose temper; angry |
| Motor Delay | motor delay; crawled at age 2, walked at 3 years; poor coordination issues; poor balance |
| Speech Delay: | Speech and language delay; struggled with talking at 3 years; receptive language delay |
| IQ | unknown |
| Intellectual Disability: | Mild intellectual disability |
| Vision: | Near- sighted, convergent squint right eye; No cortical visual impairment. He did have trouble with stairs, but this is not due to a visual issue. He had to hold on due to balance issues; constant nystagmus of the left eye when not wearing glasses |
| Hearing: | Bilateral conductive hearing loss; wears hearing aids; |
| Neurology: | Motor delay, no obvious neurological defect reported; 2021, core strength fine. |
| Neuroimaging: | No EEG or brain MRI |
| Cardiovascular: | Normal, no heart murmurs; |
| Gastrointestinal: | Feeding difficulties in infancy; Tube feed in infancy; Does independently eat now. Never any Heimlich maneuver. No choking, reflux, or vomiting. No major issues. |
| Respiratory: | Denies asthma |
| Urogenital | Undescended testicle; chordae; UTI in infancy; |
| Musculoskeletal: | Questionable leg length discrepancies; pelvis dysplasia; metatarsus adductus; short stature (2%); bilateral clinodactyly; flat occiput; pes planus sacral dimple; brachydactyly of toes; denies weak core muscles |
| Skin/Hair/Nails: | normal |
| Endocrine: | unknown |
| Immunological: | Immunodeficiency; sleep disturbance; |
| Allergies: | No known allergies |
| Metabolic: | discussion of administering growth hormone- parents opted not to |
| Neoplasm: | no |
| Inheritance: | Unknown |
| Family History of congenital disorders: | no |
| Family History of intellectual disabilities | Mother has history of epilepsy; petit mal during pregnancy-2011; |
| Consanguinity: | none |
| Medications: | No meds now |
| Services | Special Ed; Speech therapy; physical therapy starting at age 5 years; no other intervention prior to age 5. *This might affect the trajectory.* |
| Family History | Lives with both parents' and two older sisters who have no issues. |

| Survey & Parental replies to determine Eosinophil Esophagitis status: | |
|---|---|
| Question: | Answer provided by parent: |
| 1)Any allergy diagnosed? – to what? | 1) no |
| 2)Any acute skin reaction or swelling reaction to a food? | 2) no |
| 3)Anaphylaxis | 3) no |
| 4)Any episodes of diarrhea/vomiting or failure to thrive in infancy responsive to food exclusion | 4) no |
| 5)Was there any failure to gain weight? | 5) no |
| 6)Eczema – severity / extent | 6) no |
| 7) Asthma – severity / preventer | 7) no |
| 8) Allergic rhinitis | 8) no |
| 9) Odynophagia | 9) no |
| 10)Dysphagia; particularly meat and bread getting stuck. | 10) no |
| 11) Is there a need to have food cut up very small? | 11) no |

| Growth Parameters* | |
|---|---|
| Birth weight (percentile): | Wt: 2 kg. (9%) |
| Birth length: | Ht: unknown |
| Birth head circumference: | OFC: cm. (%) |
| Current weight: | Wt: 22.7 kg. (30 %) |
| Current length: | Ht: 113.4 cm. ( 1%) |
| Current head circumference: | OFC: 49.3cm. (<1%) |
| * Reference CDC Growth Charts-Simulconsult/measurement. N/A= not available | |

# H

| | |
|---|---|
| Gene Name: | ANKRD11 |
| NM_ | NM_023257_ |
| cDNA change: | c.7825C>T |
| Protein change: | p.Gln2609X |
| Genomic position: | g.16:89334035-89556969 |
| Hg build: | GRCh37 |
| Mode of inheritance: | De Novo |
| Zygosity: | heterozygous |
| Classification: | pathogenic |
| Other Variants | |
| Reason for referral | The proband is a male with developmental delays. He harbors a p.GLn2609X variant on the ANKRD11 gene diagnosed with KBG syndrome at 3 years old |

| Birth History and Vitals | |
|---|---|
| Pregnancy: | Regular prenatal care, |
| Birth (NVD or C-section): | NVD, after 4-hour labor |
| Gestational Age | 40 weeks |
| Feeding: | Feeding and swallowing difficulties in infancy, poor suck, NG-tube, and G-tube inserted, poor suck, |
| Postnatal Period: | tethered umbilical cord at birth. Cyanosis, and episodes of apnea, coughing/choking during first few days of life, failure to thrive diagnosis |

| *Facial abnormalities:* | |
|---|---|
| Ear: | |
| Eye: | |
| Nose: | Anteverted nares, |
| Mouth: | Front incisor teeth turn in. Macrodontia, very high and narrow palate, palate barely formed |
| Other: | Narrow temples, high cheek bones, craniofacial dysmorphia, |

| | |
|---|---|
| Developmental Behavior | Delayed development. Delayed milestones include first words at 19 months, crawled late at 9 months, started walking at 18 months. Anxiety, autism diagnosed, ADHD, ADD, aggression, violent behaviors, anger outburst, hitting, throwing objects, |

| | |
|---|---|
| | Was punching holes in wall prior to risperidone. Contained aggression now; handling frustrations and understanding of friendships at 4-year-old level |
| Motor Delay | Frequent falls, |
| Speech Delay: | Speech & language delay |
| IQ | unknown |
| Intellectual Disability: | Mild cognitive impairment and mild intellectual disability; functions academically below grade level in reading, writing and math |
| Vision: | hyperopia, corrected with eyeglasses. |
| Hearing: | Unilateral hearing loss, |
| ENT: | Sinus canals/cavities and ear canals malformed, recurrent sinus and ear infections |
| Neurology: | many staring spells, but not documented ever. Not clear if any other seizures since 18 months. Normal EEG. |
| Seizures/ Date of onset: | Seizure at 18 months. Grand mal seizure, no fever or infection. |
| Neuroimaging: | Normal EEG.2012, MRS and MRS of brain- normal |
| Cardiovascular: | EKG- normal, no heart issues |
| Gastrointestinal: | Feeding difficulties, swallowing difficulties and GERD, malrotation of intestines. NG tube used as infancy at 9 months, and then on to G-tube for about 1.5 years, until age 3 year. Feeds normally but has a poor appetite and food intolerance now. Does eat rapidly. Bowel control and urgency issues, |
| Respiratory: | |
| Urogenital | |
| Musculoskeletal: | Microcephaly, short stature, dolichocephaly, Extensive Wormian bones, Swollen spot on toe, due to question of bone infection, received a bone graft, with CRMO. Chronic recurring osteomyelitis treated with medication, NOT a bony spur there; small hands |
| Skin/Hair/Nails: | Abnormal hair whorl, |
| Endocrine: | Growth hormone treatment, growth hormone production found to be normal |
| Immunological: | |
| Allergies: | Food color dyes |
| Metabolic: | Failure to thrive at 9 months, Surgery for cholestoma, sleep disturbance, |
| Neoplasm: | no |
| Ob/Gyn: | unrelated mild bleeding at 12 weeks, otherwise unremarkable |
| Family History of congenital disorders: | no |
| Family History of intellectual disabilities | The oldest brother has possible Asperger's syndrome. |
| Consanguinity: | no |
| Medications: | Adderall for ADHD, which helps. Risperidone for mood control, guanfacine for ADD, growth hormones, |
| Services | 2014, IEP, Special education, OT and Speech therapies. |
| Family History | The proband lives with his biological parents, an older brother- with possible Asperger's syndrome, and a younger brother with no issues. |

| Growth Parameters* | |
|---|---|
| Birth weight (percentile): | Wt: 3.3kg. (%) |
| Birth length: | Ht: cm. (%) |
| Birth head circumference: | OFC: cm. (%) |
| Current weight: | Wt: kg. ( %) |
| Current length: | Ht: cm. ( %) |
| Current head circumference: | OFC: cm. ( %) |
| * Reference CDC Growth Charts-Simulconsult/measurement. N/A= not available | |

| HPO terms not available on website: |
|---|

Standardized Testing:

I

| | |
|---|---|
| Gene Name: | ANKRD11 |
| NM_ | NM_013275.5 |
| cDNA change: | c.7607G>A |
| Protein change: | p.Arg2536Gln |
| Genomic position: | g. |
| Hg build: | GRch37 |
| Mode of inheritance: | De novo |
| Zygosity: | Heterozygous |
| Classification: | De novo |
| Other variants: | mother's mutation, PRRT4, is associated with strong migraines with aura. Actually, she suffers such migraines, and this has become a serious problem for her. |
| Reason for referral | Proband is a 4 year 11-month-old male with Developmental Delay, and motor delay. High functioning with an estimate IQ 70-100. Family banked umbilical stem cells: Tested negative for Down syndrome. He carries mutation Arg2536Gln missense de novo - not present in mother or father. |

| Birth History and Vitals | |
|---|---|
| Pregnancy: | Normal, no alcohol nor drug consumption during pregnancy |
| Birth (NVD or C-section): | NVD |
| Gestational Age | 37 weeks |
| Feeding: | No issues |
| Postnatal Period: | No issues |

| *Facial abnormalities: facial features are like Cornelia de Lange* | |
|---|---|
| Ear: | Anteverted ears |
| Eye: | Synophrys (unibrow), Hypertelorism, thick eyebrows, prominent eyelashes, large eyes |
| Nose: | Anteverted nares, wide nasal bridge, prominent nasal tip, |
| Mouth: | Macrodontia, slightly tented upper lip vermillion, full lower lip vermillion |
| Other: | Unibrow, triangular face, short neck with low implantation of hair, Cornelia de Lange- like facial features |

| | |
|---|---|
| Developmental Behavior | developmental delay. Cannot count to 10, delay with drawing, able to write his name, but cannot copy things. Very social with other children, makes good eye contact, functions like a 2-year-old, clear delays when compared to peers. Toilet trained. Can brush his own teeth. Can put on |

| | |
|---|---|
| | some clothes, but not all of them. Trouble with pullovers. Can use fork to feed self. |
| Motor Delay: | Motor delay, low muscle tone; muscular hypotonia of the trunk |
| Speech Delay: | Language delay |
| IQ | Unknown |
| Intellectual Disability: | Developmental and cognitive delay with mild intellectual disability |
| Vision: | Wears eyeglasses, strabismus, astigmatism, no cortical visual impairment or problems with depth perception |
| Hearing: | Chronic otitis media |
| Neurology: | Hypotonia in infancy, no Babinski reflex, Episode of unrelated paroxysmal movements at 1 month of age; Delay with fontanelle closure- considering plastic helmet |
| Neuroimaging: | brain ultrasound and EEG (06/2017), normal |
| Cardiovascular: | Patent foramen ovale (now closed) |
| Gastrointestinal: | Mild problem with swallowing; some mild trouble with swallowing. One time he choked, and close to having to do Heimlich maneuver, had to clear throat with fingers. Delayed teething at 14 months, persistent primary teeth. |
| Respiratory: | None |
| Urogenital | None |
| Musculoskeletal: | Small and out-turning feet with protruding Achille's heel, brachydactyly, shortening of proximal and middle phalanges especially fifth finger, question of hip dysplasia, synovitis in hips with cold weather, fluid accumulation in the hips, hypotonia of lower extremities, joint hyperextensibility in lower limbs; short stature, delay with fontanelle closure. Soft on head, and quite delayed, with consideration for plastic helmet. |
| Skin/Hair/Nails: | None |
| Endocrine: | Never measured growth hormone. |
| Immunological: | none |
| Allergies: | None |
| Metabolic: | 2018, Blood chemistries and CBC within normal limits |
| Neoplasm: | None |
| Inheritance: | unknown |
| Family History of congenital disorders: | Maternal migraine headaches |
| Family History of intellectual disabilities | None |
| Consanguinity: | None |
| Medications: | None |
| Services | Special education attends school |
| Family History | Lives with biological parents, who were tested as a trio and do not harbor the variant. He has an older 11 year-old-sister with no developmental or health issues. Spanish is the primary language spoken in the home. |

| |
|---|
| Survey & Parental replies to determine Eosinophil Esophagitis status: |

| Question: | Answer provided by parent: |
|---|---|
| 1)Any allergy diagnosed? – to what? | 1) None |
| 2)Any acute skin reaction or swelling reaction to a food? | 2) None |
| 3)Anaphylaxis | 3) None |
| 4)Any episodes of diarrhea/vomiting or failure to thrive in infancy responsive to food exclusion | 4) None |
| 5)Was there any failure to gain weight? | 5) None |
| 6)Eczema – severity / extent | 6) None |
| 7) Asthma – severity / preventer | 7) None, episodic coughing |
| 8) Allergic rhinitis | 8) None |
| 9) Odynophagia | 9) None |
| 10)Dysphagia; particularly meat and bread getting stuck. | 10) None |
| 11) Is there a need to have food cut up very small? | 11) Not now but did have to cut up food when the boy was younger, as he did have a couple of choking episodes. No reflux |

| Growth Parameters* | |
|---|---|
| Birth weight (percentile): | 2.75kg |
| Birth length: | 46cm |
| Birth head circumference: | 22cm |
| Current weight: Nov 2021 | 18.5kg |
| Current length: Nov 2021 | 106cm |
| Current head circumference: Nov 2021 | 50cm |

| Date | weight | height | cran. per. |
|---|---|---|---|
| Nov 2020 | 15.0kg | 101cm | ? |
| Nov 2021 | 18.5kg | 106cm | 50cm |

* Reference CDC Growth Charts-Simulconsult/measurement. N/A= not available

Standardized Testing:

- Batelle Inventory: development below their chronological age in most of the areas (07/04/2017).
- CSBS: shows slight maturational delay in most areas, being greater in the motor area, expressive communication, and adaptive area (07/04/2017)

# J

| | |
|---|---|
| Gene Name: | ANKRD11 |
| NM_ | NM_013275.5 |
| cDNA change: | c. 3770_3771delAA |
| Protein change: | p. Lys1257ArgfsX25 |
| Genomic position: | g. |
| Hg build: | GRCh37 |
| Mode of inheritance: | De novo |
| Zygosity: | heterozygous |
| Classification: | pathogenic |
| Reason for referral | The proband is a 5-year-old male with global developmental delay. He carries a variant frameshift mutation p.K1257RfsX25 on the ANKRD11 gene and also CREBBP c.3393T>A variant of unknown significance. The child is macrocephaly, developmental delay, failure to thrive, partial albinism, astigmatism, facial dysmorphism, transverse palmar crease, and cryptorchidism. He received a KGB diagnosis at age 2 years and was the 78th reported person to be diagnosed with KGB syndrome. |

| Birth History and Vitals | |
|---|---|
| Pregnancy: | *Regular prenatal care, decreased movement in utero* |
| Birth (NVD or C-section): | NVD |
| Gestational Age | Term, 40 weeks |
| Feeding: | Feeding difficulty in infancy; difficulty swallowing, had tongue fasciculations; feeding tube inserted |
| Postnatal Period: | Small for age; born very short while rest of family is tall; born <1 percentile, failure to thrive at 6-week check-up; undescended testicle |

| | |
|---|---|
| Birth weight (percentile): | 3.65kg- 8lbs. 1 ounce |
| Birth length: | 17 inches- (1%) |
| Birth head circumference: | |
| Current weight: | 55% |

| | |
|---|---|
| Current height: | 110% |
| Current head circumference: | |

| | |
|---|---|
| Developmental Behavior | Global developmental delay; delayed speech and language development; tantrums; ADHD; hyperactive; No issues now with depth perception but did have past issues with going from one surface to another; sensory issues; |
| Motor Delay | Motor delay |
| Speech Delay: | Speech delay |
| IQ | unknown |
| Intellectual Disability: | yes, mild; reads at appropriate level, can identify colors and numbers up to 12, can spell name, good penmanship |
| Neurology: | Hypotonia in infancy; poor motor coordination; |
| Neuroimaging: | 2 EEGs performed, no evidence of seizures |
| *Facial abnormalities:* | Facial dysmorphia |
| Ear: | uplifted earlobe; low set ears |
| Eye: | Bilateral astigmatism |
| Nose: | broad nasal tip; depressed nasal bridge; short philtrum |
| Mouth: | Persistent primary teeth; macrodontia; thin upper lip vermillion; tongue fasciculation |
| Other: | |
| Vision: | astigmatism |
| Hearing: | Bilateral hearing loss; failed ABR; |
| Cardiovascular: | Echocardiogram done, no issues reported. |
| Gastrointestinal: | gastrotomy tube inserted in infancy- now removed, used for 3.5 years; episodic vomiting; dysphagia; tongue fasciculations |
| Respiratory: | Frequent upper respiratory infections. |
| Urogenital | cryptorchidism, urinary incontinence/dribbling |
| Musculoskeletal: | Short stature; transverse palmar crease; low muscle tone; feet curve outward; wide forehead; poor reflexes in legs for first 1 1/2 years of life, reflexes come and go, still very weak |
| Skin/Hair/Nails: | partial albinism, |

| | |
|---|---|
| Endocrine: | |
| Immunological: | Compromised immune system; |
| Allergies: | NKA |
| Metabolic: | Failure to thrive; sleep apnea; sleep disturbances; partial albinism, |
| Neoplasm: | no |
| Inheritance: | |
| Family History of congenital disorders: | no |
| Family History of intellectual disabilities | no |
| Consanguinity: | no |
| Medications: | began growth hormone injections (Humatrope) at age 3 1/2, weening off now (at time of video); no deficiency in growth hormone production; |
| Services | intense PT, speech, OT, child development. |
| Family History | Lives with parents and older sister. She has no issues. |

Standardized Testing:

# K

| | |
|---|---|
| Gene Name: | ANKRD11 |
| NM_ | NM_013275.5 |
| cDNA change: | c.1756G>A |
| Protein change: | p.Val0586Met |
| Genomic position: | g. |
| Hg build: | unknown |
| Mode of inheritance: | Maternally inherited |
| Zygosity: | Heterozygous |
| Classification: | VUS |
| Reason for referral | Multiple gene variants with global developmental delay |

| **Other Variants reported** | |
|---|---|
| Gene Name: | JMJD1C |
| NM_ | NM_032776.2 |
| cDNA change: | c.3976A>G |
| Protein change: | p.Lys1326Glu |
| Genomic position: | N/A |
| Hg build: | N/A |
| Mode of inheritance: | Maternally inherited |
| Zygosity: | Heterozygous |
| Classification: | Unknown |
| Reason for referral | Multiple gene variants with global developmental delay |

| **Other Variants reported** | |
|---|---|
| Gene Name: | TMIM71 |
| NM_ | NM_001039111.2 |
| cDNA change: | c.0707A>G |
| Protein change: | p.His0236Arg |
| Genomic position: | N/A |
| Hg build: | N/A |
| Mode of inheritance: | Autosomal Dominant |
| Zygosity: | Heterozygous (De Novo) |
| Classification: | Likely Pathogenic |
| Reason for referral | Multiple gene variants with global developmental delay |

| Birth History and Vitals | |
|---|---|
| Pregnancy: | Complicated by preeclampsia |
| Birth (NVD or C-section): | Induced early but was stuck in birth canal requiring emergent C-section |
| Gestational Age | 39 3/7 weeks |
| Feeding: | N/A |
| Postnatal Period: | Hypoxia with one minute Apgar of 1 and 5 minute 8. |

| *Facial abnormalities:* | |
|---|---|
| Ear: | Macrotia, low-set ears |
| Eye: | Synophrys, thick eyebrows, entropion in both eyes, hypertelorism |
| Nose: | Wide nasal bridge |
| Mouth: | Permanence of primary teeth: (Still has baby teeth), teeth are spaced out, thin upper lip vermilion, tented upper vermillion |
| Other: | Forward hairline, triangular face |

| | |
|---|---|
| Developmental Behavior | Developmental delay, does not know all his letters. Not as social with peers. Not entirely toilet trained and wears a pullup at night. |
| Motor Delay | Motor delays, crawled after 1 year, walked at age 27 months. Received early intervention at age 1 with PT. Problems with both gross and fine motor. Cannot peddle a bike or walk fluidly. Can feed self but has difficulty especially with spoon. |
| Speech Delay: | Speech & language delay. Speech therapy at age 2, speaks with slur, recently started to express self, syntax delay. |
| IQ | Unknown |
| Intellectual Disability: | Unknown |
| Vision: | May have astigmatism. Has hyperopia and strabismus. May have cortical visual impairment |
| Hearing: | None |
| Neurology: | Dysarthria, global developmental delay. Issues with balance with ataxic gait. Left hemiplegic cerebral palsy due to hypoxia at birth affecting balance, walking, with functional monoparesis of left hand. No hydrocephaly or headaches. Poor coordination. Has difficulty walking up and down stairs. |
| Seizures: Date of onset: | Had one seizure after surgery due to low sodium with deviated gaze only (likely simple partial seizure). Myoclonus when sleeping, unclear if seizures. |
| Neuroimaging: | MRI of brain at 4 months of age showed subdural hematoma possibly from bypass machine. No MRI of spine |
| Cardiovascular: | Tetralogy of Fallot, (TOF), with pulmonary valve sparing surgical repair at 4 months. Separated the valves, and not sure they need valve replacement. Yearly checkups, with EKG and echo. never any long QT or rhythm. Never any Holter monitor. Echo at day 8 showed VSD, aorta enlarged, pulmonary stenosis, enlarged right ventricle.<br>Surgery corrected. Ventricular Septal Defect, (VSD), and opened pulmonary valve.<br>Did not need a stent prior to surgery, as milder case of TOF. No long QT or arrythmias. Gets yearly EKGs and ECHOs. |
| Gastrointestinal: | No reflux, abdominal pain, vomiting, or choking. |
| Respiratory: | None |
| Urogenital | Cryptorchidism of right testicle requiring surgical correction, with left hydrocele that required surgery. No urinary symptoms. Insufficient foreskin with no hypospadias |
| Musculoskeletal: | Pectus carinatum. Has sacral dimple. Pronated ankles and uses SMOs and braces. Pes planus in both feet. Clinodactyly in right $5^{th}$ finger and |

| | |
|---|---|
| | all toes, no brachydactyly. Hypotonia with low core strength when younger. |
| Skin/Hair/Nails: | Hirsutism, low anterior hairline, rounded toenails |
| Endocrine: | None |
| Immunological: | None |
| Allergies: | Seasonal and amoxicillin |
| Metabolic: | Thin body built, slender, with minimal subcutaneous tissue; |
| Neoplasm: | None |
| Inheritance: | None |
| Family History of congenital disorders: | None |
| Family History of intellectual disabilities | None |
| Consanguinity: | None |
| Medications: | Claritin and Zyrtec |
| Services | Had early intervention, OT, PT, adaptive PE, speech therapy, school readiness, and IEP. Currently in virtual Headstart program. |
| Family History | Mother has scoliosis, myopia, migraines, abdominal cramping, and IBS; takes nortriptyline; no seizures. Mother has ANKRD11 gene variant. Maternal grandfather had learning disabilities in reading and math. Maternal aunt has cleft palate, learning disability, issues with kidneys, macrodontia. Brother has maternally inherited mutation missense ANKRD11. Maternal grandmother and husband are both intelligent. No consanguinity |

| Growth Parameters* | |
|---|---|
| Birth weight (percentile): | Wt: 3.72 kg (63%) |
| Birth length: | Ht: 56 cm (99%) |
| Birth head circumference: | OFC: 36 cm (<1%) |
| Current weight: | Wt: Unknown |
| Current length: | Ht: Unknown |
| Current head circumference: | OFC: Unknown |
| * Reference CDC Growth Charts-Simulconsult/measurement. N/A= not available | |

Standardized Testing: none performed for this study

# L

| | |
|---|---|
| Gene Name: | ANKRD11 |
| NM_ | NM_013275.5 |
| cDNA change: | c.1756G>A |
| Protein change: | p.Val0586Met |
| Genomic position: | g. |
| Hg build: | Unknown |
| Mode of inheritance: | Maternally inherited |
| Zygosity: | Heterozygous |
| Classification: | VUS |
| Other Variants | Unknown |
| Reason for referral | Genetically confirmed KBG syndrome, global developmental delays. Brother and mother with ANKRD11 gene variants |

| | |
|---|---|
| Gene Name: | JMJD1C |
| NM_ | NM_032776.2 |
| cDNA change: | c.3976A>G |
| Protein change: | p.Lys1326Glu |
| Genomic position: | N/A |
| Hg build: | N/A |
| Mode of inheritance: | Autosomal Dominant |
| Zygosity: | Heterozygous (Maternal) |
| Classification: | Unknown |

| Birth History and Vitals | |
|---|---|
| Pregnancy: | High risk induced 6 weeks early due to preeclampsia. No toxins or substance use |
| Birth (NVD or C-section): | Emergency C-section |
| Gestational Age | 34 weeks |
| Feeding: | Did not eat and required infantile nasogastric tube |
| Postnatal Period: | Spent two weeks in NICU for further maturation and feeds, had one episode of apnea and bradycardia |

| *Facial abnormalities:* | |
|---|---|
| Ear: | None |
| Eye: | Long eyelashes |
| Nose: | Wide nasal bridge, wide nasal base, broad nasal tip |
| Mouth: | Still has baby teeth, thin upper lip vermilion, smooth philtrum, absent Cupid’s bow. No issues with palate. |
| Other: | Broad forehead |

| | |
|---|---|
| Developmental Behavior | Global developmental delay. In a diaper now. Only follows simple commands. Very social, makes eye contact. No other evidence of autistic traits |
| Motor Delay | Delayed gross motor development. Did not crawl until 12 months, dragged one leg, can crawl normally now. Walks 3 feet before losing balance |
| Speech Delay: | Delayed speech and language development. Not talking until age 2, still babbles today and does not use words |
| IQ | Unknown |
| Intellectual Disability: | N/A |
| Vision: | N/A |
| Hearing: | Never tested |
| ENT: | Recurrent nasal congestion and sinus infections requiring antibiotics. No ear infections or tubes |
| Neurology: | Truncal hypotonia, poor coordination. No pain insensitivity, nystagmus, migraines, or headaches |
| Seizures/ Date of onset: | None |
| Neuroimaging: | None |
| Cardiovascular: | None, never did echo. Had prenatal scan at 20 weeks, recommended to do an echo. Extremities will get cold |
| Gastrointestinal: | No dysphagia can tolerate formula. No reflux or constipation |
| Respiratory: | N/A |
| Urogenital | N/A |
| Musculoskeletal: | Pronated legs, which are rigid when walking |
| Skin/Hair/Nails: | Eczema |
| Endocrine: | No bone density scan, no fractures |
| Immunological: | Palpable lymph nodes under ears |
| Allergies: | Has allergies |
| Metabolic: | Thin body build |
| Neoplasm: | N/A |
| Ob/Gyn: | N/A |
| Family History of congenital disorders: | Maternal aunt has cleft palate |
| Family History of intellectual disabilities | None |
| Consanguinity: | No |
| Medications: | None |
| Services | PT started at 13 months, mostly virtual, twice per month for 1 hr. Speech therapy starting 2 months ago for 45 min-1hr per week. No OT |
| Family History | Mother has scoliosis, myopia, migraines, abdominal cramping, and IBS; takes nortriptyline; no seizures. Mother has ANKRD11 gene variant. Older brother has multiple gene variants (ANKRD11, JMJD1C, and TMIMY1) with global developmental delay. Maternal grandfather had learning disabilities in reading and math. Maternal aunt has cleft palate, learning disability, issues with kidneys, macrodontia. Maternal grandmother and husband are both intelligent. |

| Growth Parameters* | |
|---|---|
| Birth weight (percentile): | Wt: kg. (%) |
| Birth length: | Ht: cm. (%) |
| Birth head circumference: | OFC: cm. (%) |
| Current weight: | Wt: 11.8 kg (20%) |
| Current length: | Ht: cm. (95%) |
| Current head circumference: | OFC: cm. ( %) |
| * Reference CDC Growth Charts-Simulconsult/measurement. N/A= not available | |

| HPO terms not available on website: |
|---|
| Neonatal apnea and bradycardia, poor appetite in neonate, cold extremities |

Standardized Testing: none performed for this study

# M

| | |
|---|---|
| Gene Name: | ANKRD11 |
| NM_ | NM_013275.5 |
| cDNA change: | c.1756G>A |
| Protein change: | p.Val0586Met |
| Genomic position: | g. |
| Hg build: | Unknown |
| Mode of inheritance: | Unknown |
| Zygosity: | Heterozygous |
| Classification: | Unknown |
| Other Variants | JMJD1C |
| Reason for referral | Genetically confirmed genetic variant with two sons and possibly sister with ANRD11 variants |

| | |
|---|---|
| Gene Name: | JMJD1C |
| NM_ | NM_032776.2 |
| cDNA change: | c.3976A>G |
| Protein change: | p.Lys1326Glu |
| Genomic position: | N/A |
| Hg build: | N/A |
| Mode of inheritance: | Autosomal Dominant |
| Zygosity: | Heterozygous (Maternal) |
| Classification: | Unknown |

| Birth History and Vitals | |
|---|---|
| Pregnancy: | Regular prenatal care |
| Birth (NVD or C-section): | NVD |
| Gestational Age | term |
| Feeding: | Unknown |
| Postnatal Period: | Unknown |

| *Facial abnormalities:* | |
|---|---|
| Ear: | None |
| Eye: | Thick eyebrows, no synophyrs |
| Nose: | Anteverted nares, broad nasal base |
| Mouth: | No macrodontia, smooth philtrum, or thin lip vermilion |
| Other: | None |

| | |
|---|---|
| Developmental Behavior | No cognitive issues, anxiety, currently employed |
| Motor Delay | None |
| Speech Delay: | None |
| IQ | unknown |
| Intellectual Disability: | None |
| Vision: | Myopia, eye floaters associated with pars planitis |
| Hearing: | None |
| ENT: | None |
| Neurology: | Migraines for the past 2 years associated with left sided TMJ,due to clenching for the past 3 years, numbness in left hand and foot, pain in left wrist. Right sided myokymia associated with stress |
| Seizures/ Date of onset: | No seizures |
| Neuroimaging: | None |
| Cardiovascular: | None |
| Gastrointestinal: | IBS, which started in 5$^{th}$ grade, severe to point of near syncopal episodes, and will fall to floor in pain, localizes pain to "intestines". Will have explosive diarrhea with burning pain and abdominal cramping that resolves after bowel movement, no hematochezia or melena. Could be abdominal migraines instead of IBS. No vomiting or reflux. No endoscopies or colonoscopies. |
| Respiratory: | None |
| Urogenital | None |
| Musculoskeletal: | Mild scoliosis not requiring a brace, pain in ankles, back. No swelling in joints or arthritis diagnosis |
| Skin/Hair/Nails: | None |
| Endocrine: | No swelling in joints, no arthritis diagnosis, never had growth hormone |
| Immunological: | None |
| Allergies: | None |
| Metabolic: | None |
| Neoplasm: | None |
| Ob/Gyn: | None |
| Family History of congenital disorders: | Sister has cleft palate |
| Family History of intellectual disabilities | None |
| Consanguinity: | Unknown |
| Medications: | Nortriptyline 25 mg daily for migraines and IBS |
| Services | None |
| Family History | Grandfather had a pacemaker. Father passed away from heart attack, had learning disabilities but no facial dysmorphisms. Mother had no symptoms of KBG. Sister has dyslexia, struggled academically, cleft palate, tympanostomies with recurrent otitis media, hearing problems, distinct nose, and unilateral kidney dysmorphism. Mother did not have any cardiac issues, seizures. |

| Growth Parameters* | |
|---|---|
| Birth weight (percentile): | N/A |

| | |
|---|---|
| Birth length: | N/A |
| Birth head circumference: | N/A |
| Current weight: | Normal |
| Current length: | Ht: 177.8 cm. (99%) |
| Current head circumference: | N/A |
| * Reference CDC Growth Charts-Simulconsult/measurement. N/A= not available | |

| HPO terms not available on website: |
|---|

Standardized Testing: none performed for this study

# N

| | |
|---|---|
| Gene Name: | ANKRD11 |
| NM_ | NM_013275.5 |
| cDNA change: | c.2175_2178delCAAA |
| Protein change: | p.Asn725Lysfs*23 |
| Genomic position: | g.89350777 |
| Hg build: | GRCh37 |
| Mode of inheritance: | de novo |
| Zygosity: | heterozygous |
| Classification: | pathogenic |
| Reason for referral | The proband is a 1 year 8 month old male with failure to thrive status and carries a p.Asn725LysfsX23, autosomal dominant, pathogenic variant in ANKRD11 associated with KBG syndrome |

| Birth History and Vitals | |
|---|---|
| Pregnancy: | Normal prenatal care; G3P3, GBS neg, R nonimmune, GC/C/HIV neg, CF carrier (mom and dad), APGAR 8 and 9, no drug or alcohol use during pregnancy |
| Birth (NVD or C-section): | Caesarian section, breech presentation |
| Gestational Age | 39 weeks, 5 days |
| Feeding: | Feeding difficulties, lactation consult for uncoordinated sucking; GI specialist considered NG tube, but parents increased feeding and low weight slightly improved |
| Postnatal Period: | failure to thrive, dropped to negative weight percentile at 2 months; no hypoxia, low muscle tone, floppy baby, tongue tied (corrected) |

| | |
|---|---|
| Developmental Behavior | Global developmental delay, says “mama” & “dada” appropriately, began using sign language and can sign 5-7 words, grunts and points, vocalizes without words; stranger anxiety towards adults, reluctant to maintain eye contact, good with other children; cries with rough housing. The proband is inquisitive and shows appropriate visual attention to tasks. Milestones include commenced with commando crawl at 10 months then crawled at 14 months, started walking at 18 months. Delay with sitting independently until 12 months. Could sit unsupported at 15 months. |
| Motor Delay | Motor delay. low muscle tone: arms/legs are hypertonic, torso/head are hypotonic; took 4-5 months to lift head; good finger/grip strength; delayed crawling and walking; delayed sitting up |
| Speech Delay: | delayed speech & language development, does not currently talk |
| IQ | unknown |
| Intellectual Disability: | unknown |
| Neurology: | Mild global hypotonia, hypermobility of knees & ankles, gait imbalance issues, |
| Neuroimaging: | Neuro MRI, normal per mother |
| Seizures | Never any seizures. |
| *Mild Facial dysmorphia* | |
| Ear: | Protruding ears, point outwards |
| Eye: | none |
| Nose: | Anteverted nares, broad nasal bridge, smaller nose |
| Mouth: | Macrodontia, ankyloglossia, (tongue-tied), thin lingual frenulumon with anterior attachment, high palate, arched palate, retrognathia, micrognathia, recessed mandible |
| Other: | narrow, triangular pointed chin |
| Vision: | none |
| Hearing: | none |
| Cardiovascular: | Bicuspid aortic valve and mildly dilated aortic sinuses per 2020 pediatric cardiology assessment; 2019-bilateral SVC, aortic valve tri-leaflet. foramen ovale took longer to close, systolic murmur @ left sternal edge |
| Gastrointestinal: | Feeding difficulties in infancy, poor suck, recurrent candida stomatitis, hemorrhage of anus and rectum, rectal prolapse, |
| Respiratory: | |
| Urogenital | normal male, circumcised, testes descended bilaterally |

| | |
|---|---|
| Musculoskeletal: | short stature, macrocephaly, wide rib cage, broad chest with short legs, slight lateral curving of fingers, Hip ultrasound reported normal. |
| Skin/Hair/Nails: | pilaris keratosis on legs and arms |
| Endocrine: | unknown |
| Immunological: | Possible milk allergy |
| Allergies: | none |
| Metabolic: | Failure to thrive, underweight, slender built, |
| Neoplasm: | no |
| Inheritance: | De novo |
| Family History of congenital disorders: | Yes, both parents are cystic fibrosis (CF), carriers |
| Family History of intellectual disabilities | no |
| Consanguinity: | none |
| Medications: | nystatin 100,000 unit/Ml; cholecalciferol 400U |
| Services | early intervention starting at 6 months of age, speech therapy; physical therapy |
| Family History | two older brothers: ages 4 and 6 years with no developmental issues, not tested for KBG syndrome |

| | |
|---|---|
| Birth weight (percentile): | 3225 g. (7lbs 2oz) |
| Birth length: | |
| Birth head circumference: | |
| Current weight: | 20.75 lbs (8th percentile) |
| Current length: | 31.25 in (11th percentile) |
| Current head circumference: | 49.5 cm (94th percentile) |

Standardized Testing: PDF documents available in clinical records upon request

O

| | |
|---|---|
| Gene Name: | ANKRD11 |
| NM_ | NM_013275.5 |
| cDNA change: | c.7789A>T |
| Protein change: | p.Lys2597Ter |
| Genomic position: | g. |
| Hg build: | Unknown |
| Mode of inheritance: | Paternally inherited |
| Zygosity: | Heterozygous |
| Classification: | Variant, likely pathogenic |
| Other Variants | EDA (Hypohidrotic Ectodermal Dysplasia), X-linked |
| Reason for referral | The proband is a 19-year-old male with a variant K2597X on the ANKRD11 gene. The proband has developmental delays, low muscle tone, autism, ADHD, and low pain tolerance. He was diagnosed with KBG syndrome in 2014. |

| Birth History and Vitals | |
|---|---|
| Pregnancy: | Partial abruption of placenta at 30 weeks, might have contributed to proband being born small; |
| Birth (NVD or C-section): | Normal vaginal delivery |
| Gestational Age | term |
| Feeding: | Difficulty feeding in infancy due to low muscle tone; swallow test done at age 1 showing low muscle tone and sensory-based difficulty feeding; never had feeding tube; |
| Postnatal Period: | Facial dysmorphia, feeding difficulties in infancy |

| *Facial abnormalities:* |
|---|

| | |
|---|---|
| Ear: | Recurrent ear infections; permanent hole in one eardrum; other ear has tube; |
| Eye: | Synophrys, hypertelorism, long eyelashes, |
| Nose: | Wide nasal bridge; prominent nasal tip |
| Mouth: | Retrognathia; long upper lip vermillion, poor enamel; can’t open mouth widely |
| Other: | |

| | |
|---|---|
| Developmental Behavior | Mild global developmental disabilities, anxiety; autism, ADHD, cannot express feelings verbally/not “in-tune” with feelings, expresses through anger; |
| Motor Delay | Normal walking; did not crawl; went from sitting to walking at 10 months old; |
| Speech Delay: | Neuropsychiatric evaluation at 6 years old reports sensory, motor and speech and language delays; expressive language delays, |
| IQ | Unknown |
| Intellectual Disability: | Intellectually disabled, completed high school; self-contained; currently in transition program to learn life skills; employed as grocery bagger, 20 hours/week, performing well; can write name; drew clock and time correctly but missed 11; autism; ADHD; |
| Vision: | None |
| Hearing: | Bone-anchored hearing aids; Eustachian tube was not correct size and hoped he would grow into it; conductive hearing loss determined by ENT; conductive hearing impairment |
| ENT: | Recurrent ear infections; permanent hole in one eardrum; other ear has eustachian tube insertion; |
| Neurology: | Doesn’t feel pain/high pain tolerance; |
| Seizures/ Date of onset: | None |
| Neuroimaging: | EEG done in last 6 years; MRI done with normal findings |
| Cardiovascular: | Echocardiogram done with normal findings; |

| | |
|---|---|
| Gastrointestinal: | Constipation per mother; occasionally uses stool softener; |
| Respiratory: | None |
| Urogenital | None |
| Musculoskeletal: | Kyphosis, short stature Hands are short, stubby, fat; low muscle tone, always bent over, cannot sit up straight without getting tired; cannot keep shoulders back; tight hamstrings, short legs; plays tennis and soccer well; diagnosed bone scan done with normal bone density; fractured feet, dislocated kneecap; |
| Skin/Hair/Nails: | Unibrow; nail dysplasia, nails picked down, psoriasis |
| Endocrine: | Saw endocrinologist due to growth failure; began growth hormone at age 6 until age 11 with positive results; testing showed production of growth hormone, but body wasn't responding; post natal growth retardation, |
| Immunological: | Saw immunologist due to recurrent, spontaneous fevers, immune system booster; growth hormone deficiency, |
| Allergies: | No known allergies |
| Metabolic: | body temperature dysregulation due to HED (Hypohidrotic Ectodermal Dysplasia) |
| Neoplasm: | None |
| Inheritance: | Autosomal dominant |
| Family History of congenital disorders: | Father has KBG |
| Family History of intellectual disabilities | Father has KBG, learning disabilities on paternal side of family |
| Consanguinity: | No |
| Medications: | Methylphenidate, guanfacine |
| Services | OT, ST, currently in transition program for enhancing life skills; |
| Family History | Father has KBG; older brother without KBG but never tested |

| Growth Parameters* | |
|---|---|
| Birth weight (percentile): | Wt: kg. <1 (%) |
| Birth length: | Ht: cm. (%) |
| Birth head circumference: | OFC: cm. (%) |

| | |
|---|---|
| Current weight: | Wt: 68.03 kg. 45 (%) |
| Current length: | Ht: 157.48 cm. <1 ( %) |
| Current head circumference: | OFC: cm. ( %) |
| * Reference CDC Growth Charts-Simulconsult/measurement. N/A= not available | |

| HPO terms not available on website: |
|---|

Standardized Testing: PDF documents available in clinical records upon request

P

| | |
|---|---|
| Gene Name: | ANKRD11 |
| NM_ | NM_013275.5 |
| cDNA change: | c.7789A>T |
| Protein change: | p.Lys2597Ter |
| Genomic position: | g. |
| Hg build: | Unknown |
| Mode of inheritance: | unknown |
| Zygosity: | Heterozygous |
| Classification: | Variant, likely pathogenic |
| Other Variants | |
| Reason for referral | Proband is a 59-year-old male with variant K2597X on the ANKRD11 gene. He also has a son with the same inherited mutation. |

| Birth History and Vitals | |
|---|---|
| Pregnancy: | Can't recall any difficulties |
| Birth (NVD or C-section): | Unknown |
| Gestational Age | Unknown |
| Feeding: | Unsure if any feeding issues after birth; |
| Postnatal Period: | Unknown |

| *Facial abnormalities: coarse facial features* | |
|---|---|
| Ear: | Conductive hearing loss; |
| Eye: | Poor eyesight, astigmatism; wide eyebrows, full eyelashes, |
| Nose: | Prominent nasal tip, wide nasal bridge, Large nose; |
| Mouth: | Macrodontia; bad enamel and roots, several crowns, weak teeth, broke teeth about 6 times so far while chewing soft foods; |

| |
|---|
| Other: |

| | |
|---|---|
| Developmental Behavior | Global developmental delay, mild; anxiety, depression, poor attention, has difficulty paying attention to things; currently works as cashier at grocery store; socially awkward; poor eye contact; difficulty staying focused on tasks, especially those requiring multiple steps; |
| Motor Delay | Wide gait |
| Speech Delay: | Limited expressive language by psych evaluation |
| IQ | Unknown, (IQ estimated in low-normal range by psych evaluation) |
| Intellectual Disability: | ADHD; intellectually disabled; dyslexia; has trouble with math; can't read books because he can't remember what he is reading, difficulty with memory began as early as age 8; |
| Vision: | Poor eyesight, developed in last 20 years; astigmatism; |
| Hearing: | Bilateral conductive hearing loss; uses hearing aids |
| ENT: | Recurrent sinus infections as child and now, treated with antibiotics; |
| Neurology: | Migraines for 25 years; significant memory impairment, immediately forgets information like weather, repeatedly asking same questions; some dementia; |
| Seizures/ Date of onset: | None |
| Neuroimaging: | Brain MRI performed for dementia, reported some brain atrophy; |
| Cardiovascular: | None as child; currently has atrial fibrillation with catheter ablations performed; |
| Gastrointestinal: | Irritable Bowel Syndrome: constipation, diarrhea, stomach aches from age 7 to present; dysphagia. Dysphagia began 20 years ago, starch is more difficult to swallow, |
| Respiratory: | Sarcoidosis; recurrent sinus infections; |
| Urogenital | Difficulty urinating due to enlarged prostate, on medication for this; frequent urination (polyuria); |
| Musculoskeletal: | Kyphosis; short stature; low weight; short and stubby hands; Schatzki ring (?), bilateral winged scapula; |
| Skin/Hair/Nails: | Pitted nails; fungus infection on toenails; |
| Endocrine: | None |
| Immunological: | Rheumatoid arthritis and psoriatic arthritis |
| Allergies: | No known allergies |

| | |
|---|---|
| Metabolic: | None |
| Neoplasm: | None |
| Inheritance: | |
| Family History of congenital disorders: | yes |
| Family History of intellectual disabilities | yes |
| Consanguinity: | No |
| Medications: | Humira; Methotrexate |
| Services | Attended special school as a child; |
| Family History | The subject lives with his wife of 25 years and two biological sons. One son with KBG Syndrome, another son without KBG but never tested; |

| Growth Parameters* | |
|---|---|
| Birth weight (percentile): | Wt: kg. (%) |
| Birth length: | Ht: cm. (%) |
| Birth head circumference: | OFC: cm. (%) |
| Current weight: | Wt: 58.97 kg. ( %) |
| Current length: | Ht: 157.48 cm. ( %) |
| Current head circumference: | OFC: cm. ( %) |
| * Reference CDC Growth Charts-Simulconsult/measurement. N/A= not available | |

Standardized Testing: none performed for this study

# Q

| | |
|---|---|
| Gene Name: | ANKRD11 |
| NM_ | NM_013275.5 |
| cDNA change: | c.[2273dup]: [=] |
| Protein change: | p.Arg759Glufs*23 |
| Genomic position: | g. |
| Hg build: | GRCh37 |
| Mode of inheritance: | De Novo |
| Zygosity: | heterozygous |
| Classification: | |
| Reason for referral | The proband is a 12 year old female with p.Arg759Glufs*23 variant on the ANKRD11 gene and has intellectual and developmental delays. |

| | |
|---|---|
| Birth History and Vitals | |
| Pregnancy: | very sick during pregnancy; told she might have a trisomy disorder at 16 weeks pregnant, but follow-up testing was normal |
| Birth (NVD or C-section): | vaginal delivery |
| Gestational Age | 39 weeks |
| Feeding: | difficulty feeding, milk coming out of nose, reflux; did not need feeding tube |
| Postnatal Period: | failure to thrive, apathetic baby |

| | |
|---|---|
| Developmental Behavior | Developmental delays, very learning disabled, failure to thrive, ADHD, autistic behaviors, biting, aggressive behaviors, temper tantrums, dyslexia, but did write her name, time and clock on paper; 2nd percentile in following concepts and direction; several times throughout day she seems vacant or absent/unresponsive (might be related to hearing loss); oppositional defiant disorder, gets angry and threatens with knife, pulling door off hinges |
| Motor Delay | Motor delay, floppy baby at birth, hypotonia |
| Speech Delay: | Speech & language delayed, 1st percentile in verbal reasoning; 9th percentile in decoding |
| IQ | unknown; Wechsler intelligence testing performed (low score) |
| Intellectual Disability: | intellectually impaired; attends special school; reading age 6; school doesn't think she will surpass 4th grade, has made 2 years of progress in 7 years of schooling; forgets things she has learned already; has ADHD, dyslexia, dyscalculia |
| Neurology: | fontanels didn't close until 5 or 6 years old |
| Neuroimaging: | one EEG done in June 2018, results normal |

| | |
|---|---|
| *Seizures/ onset dates:* | Dec. 2019, febrile seizures, spiked high fevers and convulsions as infant often; febrile seizure/convulsion. Some question of absence seizures (video notes) |
| *Facial abnormalities:* | Mild facial dysmorphia |
| Ear: | Normal morphology |
| Eye: | Horizontal eyebrows, bilateral amblyopia. |
| Nose: | Prominent nasal tip, broad nasal bridge upturned nose; broad nasal bridge |
| Mouth: | submucosal cleft; hard palate; long flat philtrum; macrodontia; missing several teeth and teeth are crooked; large baby teeth, bifid uvula (from genetic report) |
| Other: | single palmar crease; |
| Vision: | wears glasses; myopia (very short sighted); discrepancy between two eyes, used to have one eye patched for years; visually impaired, cannot ever drive car; eyes at -8 and -4 |
| Hearing: | Bilateral hearing loss with conductive hearing loss; received hearing aids early on; chronic ear infections; chronic pseudomonas aeruginosa infections; currently uses BAHA (bone-anchored hearing aid) |
| ENT | narrow ear canals, |
| Cardiovascular: | persistent left superior vena cava ; no ASD; normal heart |
| Gastrointestinal: | Episodic vomiting with any change from supine to sitting upright position as infant; possible sphincter control issues, No feeding tubes needed. |
| Respiratory: | possibly had COVID or unknown virus in December 2019; history of severe bronchiolitis at 12 days old; history of pneumonia |
| Urogenital | urinates herself during febrile convulsions |
| Musculoskeletal: | Short stature, clinodactyly, short fingers, lateral curving, coordination issues with fingers; toe syndactyly (webbed toes), pes planus wide/flat feet; curved spine, sits hunched over, Scheuermann's kyphosis; can barely touch toes, chronic leg and foot pain, tight hamstrings contractures, |
| Skin/Hair/Nails: | |
| Endocrine: | precocious puberty? starting at age 9, and periods around age 11. whereas mother was 15 at her puberty. |
| Immunological: | chronic ear infections, chronic pseudomonas aeruginosa infections, recurrent infections |
| Allergies: | |
| Metabolic: | Sleep disturbance (takes melatonin), failure to thrive, cannot regulate or control body temperature, (always hot), high pain threshold, |
| Neoplasm: | no |
| Inheritance: | Autosomal recessive (in absence of a haploid insufficiency choice) |
| Family History of congenital disorders: | no |

| | |
|---|---|
| Family History of intellectual disabilities | no |
| Consanguinity: | no |
| Medications: | melatonin |
| Services | Special Ed classes, |
| Family History | Lives with mother, parents divorced, 14-year-old brother and 8-year-old sister with no developmental issues; not tested for KBG syndrome |

**Growth Parameters:**

| | |
|---|---|
| Birth weight (percentile): | 3.26 kg 7 lbs 3 oz (<1 percentile) |
| Birth length: | NA |
| Birth head circumference: | NA |
| Current weight: | NA |
| Current length: | NA |
| Current head circumference (OFC): | NA |

Standardized Testing: none performed for this study

R

| | |
|---|---|
| Gene Name: | ANKRD11 |
| NM_ | NM_031275.5 |
| cDNA change: | c.6596_6597insA |
| Protein change: | p.Ala2201CysfsTer |
| Genomic position: | g. |
| Hg build: | N/A |
| Mode of inheritance: | De novo |
| Zygosity: | Heterozygous |
| Classification: | Pathogenic |
| Other Variants | N/A |
| Reason for referral | The proband is a 20-year-old female who is developmentally delayed and has a Ala2201CysfsTer variant on the ANKRD11 gene |

| Birth History and Vitals | |
|---|---|
| Pregnancy: | Bleeding at 16 weeks, went to hospital. Choroid plexus cysts on brain at 20 weeks (suspicion of Edwards syndrome), resolved one month later. Contractions at 33 weeks stopped with morphine. One week later, heart rate decelerations from 150 to 30 and had emergency C-section at 34 weeks |
| Birth (NVD or C-section): | Emergent C-section |
| Gestational Age | 34 weeks |
| Feeding: | Difficulty with feeding, poor suck reflex |
| Postnatal Period: | Stayed in hospital for 12 days, did not need ventilation. Mother noticed eyes closed, very white, no hair. High pitched crying, rocking, holding self upside-down |

| *Facial abnormalities:* | |
|---|---|
| Ear: | Posterior dimples, attached ear lobes |
| Eye: | Synophrys, epicanthal folds, almond shaped eyes, large eyebrows, hypertelorism |
| Nose: | Anteverted nares, wide nasal bridge, broad nasal base, button nose when younger but now broad nose tip |
| Mouth: | Macrodontia (wide teeth), never had cavities. Had arthrocentesis due to inability to open mouth possibly due to arthritis, which made eating difficult. V-shaped and narrow jaw, jaw protruding with underbite. |
| Other: | Triangular face, prominent forehead |

| | |
|---|---|
| Developmental Behavior | Global developmental delays finished secondary school. Saw psychologist for behaviors. Has learning disability but not specified. Passed exams for French and Spanish. Poor in math, English (memory issue), and program solving. Cognitive and emotional level of a 13-year-old. Rhythmic screaming, frequently with temper tantrums with |

| | |
|---|---|
| | one episode lasting 23 hours, did not resolve with melatonin. Would hit mouth and ear for hours, head banging. Behaviors worsen with menses. Diagnosed with autism at age 4. Has sensory issues with certain textures, does not like things that are wet or sticky. Delay with toilet training until 4 years. |
| Motor Delay | Motor delays, Delay with walking, would crawl on head |
| Speech Delay: | Only two words at age 2.5 |
| IQ | N/A |
| Intellectual Disability: | Yes, diagnosed at 17 years |
| Vision: | Wears glasses, strabismus of right eye, |
| Hearing: | Decreased hearing until age 2, failed tests until age 2. May still have issues in left ear, difficulty with low frequency |
| ENT: | No sinus infections but often had ear infections, no ear tubes |
| Neurology: | Had LP in the past (possible swelling in spine?), no migraines; has strabismus of right eye. Decreased pain sensation. Walks with wide gait and has coordination issues |
| Seizures/ Date of onset: | Had seizure at a few days of age (stopped breathing) with subsequent “floppy” episode, absence seizures during school. Developed tonic-clonic seizures at 16, hit head during first episode, and went to hospital, had a repeat seizure one month later. Diagnosed with EEG, now on lamotrigine. |
| Neuroimaging: | MRI at age 4 to rule out Cinca syndrome. Another MRI brain at 17 years. X-ray and MRI spine showed elongated vertebral bodies with endplate changes and loss of anterior vertebral body height |
| Cardiovascular: | Physiologic murmur when child |
| Gastrointestinal: | Constipation as an infant, took lactulose |
| Respiratory: | N/A |
| Urogenital | Frequent urinary infections and fevers as infant |
| Musculoskeletal: | Clinodactyly of 5$^{th}$ digits, brachydactyly of 4$^{th}$ and 5$^{th}$ digits as well as all toes. Swollen PIP joints on 1$^{st}$ and 2$^{nd}$ digits. Single palmar crease on right hand. No sacral dimple, delayed fontanelle closed at 2.5 years. Broad neck. Short stature compared to family |
| Skin/Hair/Nails: | Dystrophic toenails with large toenails removed. Urticaria |
| Endocrine: | Early onset of puberty at age 10, Early menses. No diagnosis of osteopenia but fractured femur at age 4. Never treated with growth hormone. |
| Immunological: | Juvenile arthritis never had bone density scan. Treated with ibuprofen, methotrexate, and steroid injections. Now on abatacept |
| Allergies: | No known allergies |
| Metabolic: | Weight remained in 2$^{nd}$ percentile, saw dietician for high calorie shakes and high calorie diet |
| Neoplasm: | no |
| Ob/Gyn: | On Cerazette to prevent menses as it is associated with worsened behavior. Menses started at age 10 |
| Family History of congenital disorders: | Younger brother has Chiari type 1 malformation |
| Family History of intellectual disabilities | no |
| Consanguinity: | no |

| | |
|---|---|
| Medications: | Risperidone for screaming. Currently on abatacept, lamotrigine, and Cerazette |
| Services | Had special ed as a child but transition to general ed, had special small group learning and extra help in high school. PT at 1.5 years, OT at 4 years, speech therapy at 3 years |
| Family History | The proband lives with her mother, the parents are divorced. The proband has two brothers (one fraternal twin who was tested and negative for KBG, and one older), older brother does not appear to have KBG. Twin brother had benign Rolandic epilepsy |

| Growth Parameters* | |
|---|---|
| Birth weight (percentile): | Wt: 2.4 kg (<1%) |
| Birth length: | Ht: 46.5 (9%) |
| Birth head circumference: | OFC: N/A |
| Current weight: | Wt: 58 kg (52%) |
| Current length: | Ht: 162.6 cm (46%) |
| Current head circumference: | OFC: N/A |
| * Reference CDC Growth Charts-Simulconsult/measurement. N/A= not available | |

Standardized Testing: none performed for this study

S

| | |
|---|---|
| Gene Name: | ANKRD11 |
| NM_ | NM_031275.5 |
| cDNA change: | c.5659C>T |
| Protein change: | p.Gln1887* |
| Genomic position: | g. |
| Hg build: | Unknown |
| Mode of inheritance: | Autosomal Dominant |
| Zygosity: | Heterozygous |
| Classification: | Pathogenic |
| Other Variants | None |
| Reason for referral | the proband is a 11-year-old girl with Attention Deficit/Hyperactivity Disorder and Specific Learning Disorder with impairment in reading. Her medical history is notable for epilepsy and KGB syndrome. |

| Birth History and Vitals | |
|---|---|
| Pregnancy: | Normal prenatal care |
| Birth (NVD or C-section): | NVD with vacuum assistance |
| Gestational Age | term |
| Feeding: | N/A |
| Postnatal Period: | N/A |

| *Facial abnormalities:* | |
|---|---|
| Ear: | None |
| Eye: | trichomegaly, synophrys, slightly up slanting palpebral fissures, long eyelashes, thick and arched eyebrows, |
| Nose: | Anteverted nares, broad nasal root with bulbous nasal tip |
| Mouth: | Macrodontia, deeply grooved philtrum with thin upper lip, narrow palate |
| Other: | None |

| | |
|---|---|
| Developmental Behavior | Global developmental delay. motor, social, and language developmental milestones were delayed, enjoys being with people, ADHD, poor impulse, and emotional control with aggressive behavior toward brother and dog, dysgraphia, dysmetria, dyslexia, delayed toilet training. Would fill mouth with food before swallowing. Negative for autism, no stereotypies, no fascinations in special topics. Unspecified anxiety disorder, unspecified obsessive-compulsive and related disorder. |

| | |
|---|---|
| Motor Delay | Delay with sitting, crawling, walking, fine motor control delay more so than gross |
| Speech Delay: | Delayed language |
| IQ | 77 |
| Intellectual Disability: | None |
| Vision: | Glasses for nearsightedness, no issues with depth perception |
| Hearing: | None |
| ENT: | N/A |
| Neurology: | unsteady gait, shuffling gait, hypotonia, sleep disturbances, sensory differences (tactile and sensory); abdominal migraines, |
| Seizures/ Date of onset: | epilepsy (myoclonic and febrile seizures to generalized tonic-clonic seizures) |
| Neuroimaging: | MRI Brain at time of epilepsy and KBG diagnoses (Multiple none enhancing T2/FLAIR hyperintense subcortical lesions in left frontal lobe 2012), frequent EEGs |
| Cardiovascular: | low exercise tolerance, no other issues, normal Echo and EKG |
| Gastrointestinal: | GERD, episodic vomiting, dysphagia, abdominal migraines, |
| Respiratory: | no asthma |
| Urogenital | diurnal enuresis |
| Musculoskeletal: | no webbed feet or hands, flat feet, feet turned inwards, gluteal cleft, negative for spina bifida; lordosis, sacral dimple, brachycephaly, bitemporal narrowing, |
| Skin/Hair/Nails: | sensitive skin, flaking skin on head |
| Endocrine: | None |
| Immunological: | None |
| Allergies: | None |
| Metabolic: | iron deficiency, sleep disturbance, |
| Neoplasm: | None |
| Inheritance: | Heterozygous, |
| Family History of congenital disorders: | None |
| Family History of intellectual disabilities | Immediate and extended family mental health history includes anxiety, depression, OCD, panic attacks, schizophrenia, and substance abuse. |
| Consanguinity: | None |
| Medications: | Lamotrigine 5 mg twice daily, methylphenidate 36 mg daily, Guanfacine, CBD 25 mg twice daily, PPI |
| Services | IEP, OT for writing, PT for walking, speech therapy |
| Family History | Father has aortic stenosis, maternal grandma with auditory schwannomas and acoustic neuromas. Anxiety, depression, OCD, panic disorder, schizophrenia, and substance use. |

| Growth Parameters* | |
|---|---|
| Birth weight (percentile): | Wt: 3.03 kg. (23%) |
| Birth length: | Ht: |
| Birth head circumference: | OFC |
| Current weight: | Wt: 37.6 kg. (1%) |
| Current length: | Ht: 129.54 cm. (<1%) |

| | |
|---|---|
| Current head circumference: | OFC: 50 cm. (<1%) |
| * Reference CDC Growth Charts-Simulconsult/measurement. N/A= not available | |

Normal fragile X testing including karyotype, chromosomal microarray, NIPSL sequencing and del/dup.

Standardized Testing: WISC-V: Full scale IQ of 77, previously 86 in 2018

T

| | |
|---|---|
| Gene Name: | ANKRD11 |
| NM_ | NM_013275.5 |
| cDNA change: | c.5227C>T |
| Protein change: | p.Gln1743Ter |
| Genomic position: | g. NA |
| Hg build: | GRch37/hg19 |
| Mode of inheritance: | Autosomal Dominant |
| Zygosity: | Heterozygous |
| Classification: | Pathogenic variant |
| Reason for referral | Individual with autism, global developmental delay, developmental regression, ADHD, intellectual disability, atrioventricular septal defect, and dysmorphic features. |

| Birth History and Vitals | |
|---|---|
| Pregnancy: | Difficult pregnancy: twin pregnancy, second child was stillborn; placenta previa; baby stopped moving during development- no obvious hypoxia; during the delivery baby's heart rate was undetectable and had to use probe |
| Birth (NVD or C-section): | NVD |
| Gestational Age | 32-34 weeks |
| Feeding: | NA |
| Postnatal Period: | NA |

| *Facial abnormalities:* | |
|---|---|
| Ear: | None |
| Eye: | Synophrys (Unibrow); Arched eyebrows |
| Nose: | Wide nasal bridge |
| Mouth: | Macrodontia; severe ankyloglossia; possible jaw deformity- lock jaw and lack of mobility; dental crowding |
| Other | Cornelia de Lange like features; Pointed chin |

| | |
|---|---|
| Developmental Behavior | Developmental delay, Socially awkward, socially anxious, does not have many friends; behavioral reactions to preservatives in food/medications (molasses, red dye, raisins, propofol, specific ADHD medication) makes him aggressive and violent |
| Motor Delay | Motor delay, delayed crawling, walking, and running; learning how to use utensils now; lack of upper body strength |
| Speech Delay: | Began talking at 3 years old |
| IQ | IQ71 |

| | |
|---|---|
| Intellectual Disability: | Global developmental delay; Autism, Disruptive Mood Dysregulation Disorder, ADHD, anxiety, intellectual delay; functioning like 4yr old at 8 years |
| Vision: | None |
| Hearing: | Bilateral otitis media at 2 yrs.; tubes placed x 5-6 times; adenoidectomy; T-tube placed in left ear due to eustachian tube collapse- hearing ok now, right ear ok; never had hearing aid |
| Neurology: | Seizures starting at age 2; staring spells and unresponsive, absence seizures; has migraines x 4 years with visual aura; sleepwalking; difficulties with sleep onset |
| Neuroimaging: | None |
| Seizures/onset: | absence seizures; |
| Cardiovascular: | Congenital heart defect detected at 1 month; open heart surgery at 1 year (9/28/12); Atrial septal defect; mitral valve repair with mild insufficiency currently; atrial ventricular defect; heart rhythm issues |
| Gastrointestinal: | Abdominal migraines; cramping, vomiting, diarrhea; constipation and gas; acid reflux; no feeding tube |
| Respiratory: | Asthma, pulmonary edema, |
| Urogenital | None |
| Musculoskeletal: | Generalized hypotonia; hypotonia upper and lower extremities; normal hands- no obvious clinodactyly, no obvious single palmar crease; flat feet; no sacral dimple; no bone issues |
| Skin/Hair/Nails: | None |
| Endocrine: | unspecified aplastic anemia; blood transfusions with high reticulocyte count |
| Immunological: | None |
| Allergies: | allergy to antibiotic medication |
| Metabolic: | None |
| Neoplasm: | None |
| Inheritance: | Mother is mosaic for mutation |
| Family History of congenital disorders: | Stillborn twin birth. |
| Family History of intellectual disabilities | None |
| Consanguinity: | NA |
| Medications: | Focalin XR; amantadine; Allegra; Singular; Flonaze; pantoprazole; magnesium; VitB2; Zomig; Sennalax |
| Services | Special education; speech and physical therapy starting at 7 years; Applied Behavior Analysis therapy |
| Family History | Mother had miscarriage last April; mother has copper toxicity (ruled out Wilsons in August); mother has anemia; mother has history of seizure as a child; mother has problems with sensory processing |

| | |
|---|---|
| Survey & Parental replies to determine Eosinophil Esophagitis status: | |
| Question: | Answer provided by parent: |
| 1)Any allergy diagnosed? – to what? | 1) antibiotics |

| | |
|---|---|
| 2)Any acute skin reaction or swelling reaction to a food? | 2) None |
| 3)Anaphylaxis | 3) None |
| 4)Any episodes of diarrhea/vomiting or failure to thrive in infancy responsive to food exclusion | 4) gastric migraines |
| 5)Was there any failure to gain weight? | 5) None |
| 6)Eczema – severity / extent | 6) None |
| 7) Asthma – severity / preventer | 7) yes, asthma |
| 8) Allergic rhinitis | 8) no |
| 9) Odynophagia | 9) |
| 10)Dysphagia; particularly meat and bread getting stuck. | 10) |
| 11) Is there a need to have food cut up very small? | 11) |

| Growth Parameters* | |
|---|---|
| Birth weight (percentile): | Wt: 3.4kg. (16.0%) |
| Birth length: | Ht: 49.5cm. (10.4%) |
| Birth head circumference: | OFC- |
| Current weight: | Wt- |
| Current length: | Ht- |
| Current head circumference: | OFC- |
| * Reference CDC Growth Charts-Simulconsult/measurement. N/A= not available | |

4/1/14:
Chest Radiograph- cardiomegaly with changes suggestive of pulmonary edema
CT Abdomen/Pelvis without contrast: bilateral pulmonary edema, small amount of fluid in lower pelvis

5/2/14:
Echocardiogram- history of partial AV canal defect s/p surgical repair on 9/29/12 with good result; top normal velocities across mitral valve with trivial-mild insufficiency, normal biventricular dimensions and systolic function

**Standardized Testing:**

7/15/2019:

**Reynolds Intellectual Assessment Scales**
Verbal Intelligence Index (VIX) score: 83 (lower 1/3 of low average range)
Nonverbal Intelligence Index (NIX): 66 (intellectually deficient)
Composite Intelligence Index (CIX): 71 (below average)
Composite Memory Index (CMX): 75 (below average)
**Vineland-3 Adaptive Behavior Scales:** reveals Joshua is functioning below age-level expectations in socialization, communication and motor skills
**Autism Composite Score:** 75 (high probability of autism)
**ADOS-2 Score:** 5 (moderate)

# U

| | |
|---|---|
| Gene Name: | ANKRD11 |
| NM_ | NM_013275.5 |
| cDNA change: | c.5227C>T |
| Protein change: | p.Gln1743Ter |
| Genomic position: | g. NA |
| Hg build: | GRch37/hg19 |
| Mode of inheritance: | unknown |
| Zygosity: | Heterozygous |
| Classification: | Pathogenic variant |
| Other Variants | |
| Reason for referral | **Low level of mosaicism for ANKRD11 tested by saliva;** son with ANKRD11 mutation |

| | |
|---|---|
| Birth History and Vitals | |
| Pregnancy: | Born at 4lb 3oz |
| Birth (NVD or C-section): | NVD |
| Gestational Age | Full-term |
| Feeding: | Underweight until 14 years |
| Postnatal Period: | Did not eat a lot; small stature in infancy |

| | |
|---|---|
| *Facial abnormalities:* | |
| Ear: | None |
| Eye: | Synophrys, thick eyebrows |
| Nose: | Wide nasal bridge, prominent nasal tip |
| Mouth: | Root canal; carious teeth, yellow-brown discoloration of the teeth; macrodontia |
| Other: | |

| | |
|---|---|
| Developmental Behavior | ADHD; finished some college; short attention span, sensory adversity towards Styrofoam |
| Motor Delay | Frequent falls, fell a lot especially going up stairs; clumsy; poor motor coordination; bump into walls; problems with depth perception |
| Speech Delay: | Language delay, received speech therapy |
| IQ | unknown |
| Intellectual Disability: | unknown |
| Vision: | blurred vision |
| Hearing: | None |

| | |
|---|---|
| ENT: | Left sinus repair and septum surgery 2020; deviated septum; sinus infections; sinus infection spread to gums; otitis externa x3; loss of taste and smell, esophageal spasms (COVID); tonsillectomy; wisdom teeth removal; multiple cavities filled |
| Neurology: | Migraines with aura; hospitalized for migraines and vomiting at age 17 and was given migraine cocktail with antinausea meds; history of fainting; narcoleptic fits (covid effect) |
| Seizures/ Date of onset: | seizure at 5 years of age |
| Neuroimaging: | Brain MRI done |
| Cardiovascular: | Baseline heart rate ~118; now at 102 bpm; tachycardia; bradycardia (during covid) |
| Gastrointestinal: | Abdominal migraines, nausea, vomiting; gut spasms; constipation; blood in stool; loss of appetite (COVID effects); weight loss 90lbs in past year; Irritable bowel syndrome; acid reflux |
| Respiratory: | Trouble breathing due to reported COVID infection on 4/12/21; chest XR normal |
| Urogenital | Pollakisuria, frequent urination; congenital knots in urethra; spasms in pelvic area; urethra collapsed and had to be dilated; overactive bladder |
| Musculoskeletal: | Fell and broke hand and arm resulting in chronic pain during military training; bone density scan revealed hypocalcemia and potential for osteoporosis; short stature in infancy |
| Skin/Hair/Nails: | None |
| Endocrine: | High levels of copper toxicity in blood (ruled out Wilsons); problems with potassium levels; low B12; high urea nitrogen levels |
| Immunological: | None |
| Allergies: | None |
| Metabolic: | Slender built, |
| Neoplasm: | None |
| Inheritance: | Autosomal Dominant |
| Family History of congenital disorders: | Son diagnosed with KBG mutation |
| Family History of intellectual disabilities | Son also diagnosed with autism, ADHD, global developmental delay, intellectual disability |
| Consanguinity: | NA |
| Medications: | Hyoscyamine, Mirabegron and another medication for bladder, Galcanezumab stopping and switching to Ajovy for migraines, Tylenol, Ibuprofen, Calcium, B12, Potassium, Multivitamin with Iron, plecanatide for IBS, dicyclomine for IBS, Zofran, Zyrtec, biotin, pantoprazole, Maxalt, Adderall |
| Services | Speech therapy as a child |
| Family History | History of 2 miscarriages; mother, father, and son have migraines; brother has speech impediments and a lisp |

| Growth Parameters* | |
|---|---|
| Birth weight (percentile): | Wt: 1.90 kg. (<1 %) |
| Birth length: | Ht: cm. (%) |
| Birth head circumference: | OFC: cm. (%) |

| | |
|---|---|
| Current weight: | Wt: kg. ( %) |
| Current length: | Ht: cm. ( %) |
| Current head circumference: | OFC: cm. ( %) |
| * Reference CDC Growth Charts-Simulconsult/measurement. N/A= not available | |

| HPO terms not available on website: |
|---|
| Abdominal migraines<br>Increased circulating copper concentration |

Standardized Testing: none performed for this study

# V

| | |
|---|---|
| Gene Name: | ANKRD11 |
| NM_ | NM_001256182.1 |
| cDNA change: | c.1977C>G |
| Protein change: | p.Tyr0659* |
| Genomic position: | g. |
| Hg build: | GRCh37(Hg19) |
| Mode of inheritance: | De Novo |
| Zygosity: | Heterozygous |
| Classification: | Likely pathogenic |
| Other Variants | |
| Reason for referral | The proband is a 5-year 11 months female with motor delays wh carries polygenic variants. |

| | |
|---|---|
| Gene Name: | MUTYH |
| NM_ | NM_001128425.1 |
| cDNA change: | c.1187G>A |
| Protein change: | p.Gly0396Asp |
| Genomic position: | g. |
| Hg build: | GRCh37(Hg19) |
| Mode of inheritance: | De Novo |
| Zygosity: | Heterozygous |
| Classification: | Pathogenic |

| | |
|---|---|
| Gene Name: | SELENON |
| NM_ | NM_020451.2 |
| cDNA change: | c.0943G>A |
| Protein change: | p.Gly0315Ser |
| Genomic position: | g. |
| Hg build: | GRCh37(Hg19) |
| Mode of inheritance: | De Novo |
| Zygosity: | Heterozygous |
| Classification: | Pathogenic |

| Birth History and Vitals | |
|---|---|
| Pregnancy: | On fertility drug Femara, infant was small for gestational age |
| Birth (NVD or C-section): | NVD |
| Gestational Age | 40 weeks + 4 days, late |
| Feeding: | None |
| Postnatal Period: | Mild neonatal jaundice |

| *Facial abnormalities:* | |
|---|---|
| Ear: | None |
| Eye: | Hypertelorism, thick eyebrows |

| | |
|---|---|
| Nose: | Anteverted nares, broad nasal tip, broad nasal bridge |
| Mouth: | Has primary set of teeth, thin lip vermillion. High arched palate, ankyloglossia. No macrodontia |
| Other: | Triangular face, frontal bossing |

| | |
|---|---|
| Developmental Behavior | Global developmental delay. Able to write name, struggles with letter sequence, toilet trained, very social with good eye contact, can feed self, issues with fine motor coordination,( writing, dyscalculia, dysgraphia, ), has tantrums, (abnormal affect behavior), but does not understand punishment or cause and effect, may have difficulty with math, difficulty telling stories in chronological order, good at memorization |
| Motor Delay | Walking at 21 months, crawled at 10 months, did not play on playground until 4, delay with throwing/catching balls, climbing ladders, and tying shoes. Gross motor delay. |
| Speech Delay: | Not talking at 18 months, started at 20 months, did not babble. Expressive language disorder |
| IQ | Not tested, |
| Intellectual Disability: | Unknown, negative for Fragile X syndrome |
| Vision: | No issues, no depth perception issues |
| Hearing: | Mixed sensorineural and conductive hearing loss bilaterally, worse in right, bilateral myringotomies 2-3 times due to fluid accumulation, large ear canals, sedated ABR, has hearing aid in right |
| Neurology: | Issues with coordination, no migraines or headaches, good memory |
| Seizures/ Date of onset: | No seizures, never had EEG |
| Neuroimaging: | Brain MRI at 18 months was normal |
| Cardiovascular: | No cardiac history, never had echo |
| Gastrointestinal: | Gastrointestinal Acid reflux, projectile vomiting up to 6 months of age after feeds, mother stopped having dairy with resolution of vomiting, not tested for lactose intolerance. Had 3 instances of multiple episodes of vomiting followed by more vomiting on day 3, has not ruled out cyclic vomiting syndrome, does not appear to be abdominal migraines |
| Respiratory: | None |
| Urogenital | Ultrasound kidneys on 6/2/20 showed normal kidneys with partially distended urinary bladder |
| Musculoskeletal: | Sacral dimple, thin frame, mild clinodactyly in 5$^{th}$ left digit, hip dysplasia requiring harness for 12 weeks, hypermobile, had braces on feet, hypotonia, weak core strength, persistent open anterior fontanelle indented with severe dipping, knee pain, leg pain, dull aches without swelling. Feet were not inverted, did have pes planus. Short stature when younger, which has since resolved, no pes excavatum or carinatum, slender built. |
| Skin/Hair/Nails: | None |
| Endocrine: | Elevated TSH but normal thyroid levels |
| Immunological: | Adenoid hypertrophy, adenoidectomy, chronic pansinusitis, recurrent otitis media |
| Allergies: | None |
| Metabolic: | None |
| Neoplasm: | None |

| | |
|---|---|
| Inheritance: | None |
| Family History of congenital disorders: | None |
| Family History of intellectual disabilities | None |
| Consanguinity: | No |
| Medications: | None |
| Services | Two 30-minute OT sessions per week, 3 each 30-minute special ed services per week, one 30 minute hard of hearing per week, observation for 30 minutes. Early intervention at 18 months, PT and speech therapy until 3 years old, had special education and speech therapy for delays, IEP, class for deaf children |
| Family History | Younger sister without any delays, mother has ADHD and seasonal allergies. Paternal grandmother has autoimmune disease and died before age 50. Maternal grandfather had autoimmune disease |

| Growth Parameters* | |
|---|---|
| Birth weight (percentile): | Wt: 2.948 kg. (18%) |
| Birth length: | Ht: 49.53 cm. (51%) |
| Birth head circumference: | OFC: Unknown |
| Current weight: | Wt: 40-50% |
| Current length: | Ht: 40-50% |
| Current head circumference: | OFC: Unknown |
| * Reference CDC Growth Charts-Simulconsult/measurement. N/A= not available | |

| HPO terms not available on website: |
|---|
| Knee pain, adenoidectomy, mother with group B strep, (maternal sepsis) |

Standardized Testing: Visual Motor Integration is very low. Visual perception is below average. Motor coordination is low

# W

| | |
|---|---|
| Gene Name: | ANKRD11 |
| NM_ | NM_013275.5 |
| cDNA change: | c.2409_2412del |
| Protein change: | p.Glu805Argfs*57 |
| Genomic position: | g.89350538_89350541del |
| Hg build: | GRch37hg19 |
| Mode of inheritance: | De Novo |
| Zygosity: | Heterozygous |
| Classification: | Pathogenic |
| Reason for referral | The proband has Severe intellectual disability; KGB syndrome; myoclonic epilepsy |

| Birth History and Vitals | |
|---|---|
| Pregnancy: | No antenatal problems during pregnancy; mom was on Propranolol which she had been on since her 20s for anxiety |
| Birth (NVD or C-section): | Emergency C-section |
| Gestational Age | 40+1 weeks |
| Feeding: | Breastfed for 3 weeks then switched to mixed feeding |
| Postnatal Period: | No postnatal problems |

| | |
|---|---|
| Developmental Behavior | Severe Developmental Delays: not toilet trained; self-stimulating behaviors; Autism diagnosed in 2018; Substantial developmental regression; Absent language; obsessive behaviors; Developmental regression, |
| Motor Delay | Motor delays, Crawled at 1.5 years; Started walking at 2 years |
| Speech Delay: | Speech & language delays, Spoke 30 words total and put 2 words together until age 4; does not speak at all now. Regression of speech. |
| IQ | unknown |
| Intellectual Disability: | Severe intellectual disability; generalized developmental delay; speech and language delay; autism, autistic be; functions like 7-month-old according to mother |
| Vision: | normal |
| Hearing: | normal |
| Neurology: | Myoclonic epilepsy with drop attacks and myoclonic jerks; large prominent fontanelles |
| Neuroimaging: | MRI Head with contrast on 10/18/2016 was unremarkable with only slight prominence of extra-axial spaces frontally; EEG- myoclonic epilepsy |
| Seizures: | EEG- myoclonic epilepsy |
| Cardiovascular: | None; EKG normal |
| Gastrointestinal: | Trouble feeding as infant; considering percutaneous endoscopic gastrostomy (PEG); acid reflux starting at age 6 and now 1x/week; eats pureed food |

| | |
|---|---|
| Respiratory: | None |
| Urogenital | None |
| Musculoskeletal: | Central and mild peripheral hypotonia, small hands with clinodactyly- curving,small 5th finger, small feet with no webbing, no sacral dimple |
| Skin/Hair/Nails: | None |
| Endocrine: | None |
| Immunological: | None |
| Allergies: | None |
| Metabolic: | elevated alkaline phosphatase; probable biotinidase deficiency |
| Neoplasm: | None |
| Inheritance: | None |
| Family History of congenital disorders: | None |
| Family History of intellectual disabilities | Maternal uncle with learning disability |
| Consanguinity: | no |
| Medications: | Clobazam, Omeprazole, Furosemide, Rufinamide, Melatonin |
| Services | Attends specialist education school |
| Family History | NA |

| | |
|---|---|
| Survey & Parental replies to determine Eosinophil Esophagitis status: | |
| Question: | Answer provided by parent: |
| 1)Any allergy diagnosed? – to what? | 1) No food allergy |
| 2)Any acute skin reaction or swelling reaction to a food? | 2) No |
| 3)Anaphylaxis | 3) No |
| 4)Any episodes of diarrhea/vomiting or failure to thrive in infancy responsive to food exclusion | 4) No |
| 5)Was there any failure to gain weight? | 5) No |
| 6)Eczema – severity / extent | 6) No |
| 7) Asthma – severity / preventer | 7) No |
| 8) Allergic rhinitis | 8) No |
| 9) Odynophagia | 9) No |
| 10)Dysphagia; particularly meat and bread getting stuck. | 10) No |

| | |
|---|---|
| 11) Is there a need to have food cut up very small? | 11) Not really due to zero choking |

| Growth Parameters* | |
|---|---|
| Birth weight (percentile): | Wt: 3 kg. (5.6%) |
| Birth length: | Ht: - cm. (9%) |
| Birth head circumference: | OFC: - |
| Current weight: | Wt: 18.9kg. (5.8%) |
| Current length: | Ht: 117cm. (18 %) |
| Current head circumference: | OFC: - |
| * Reference CDC Growth Charts-Simulconsult/measurement. N/A= not available | |

Standardized Testing:
3/2014: Brain MRI- normal
3/27/2014: US abdomen- normal
2014: Renal US-normal
2014: Wrist XR- normal
12/18/2014: EEG- myoclonic epilepsy without hypsarrhythmia

# X

| | |
|---|---|
| Gene Name: | ANKRD11 |
| NM_ | NM_031275.5 |
| cDNA change: | c.1893delA |
| Protein change: | p.Lys0631fs |
| Genomic position: | g. |
| Hg build: | GRCh37 |
| Mode of inheritance: | De Novo |
| Zygosity: | Heterozygous |
| Classification: | Pathogenic |
| Second variant | Heterozygous ARHGEF9 c.1496G>A p.Arg499His; reported to cause X- linked recessive Early infantile epileptic encephalopathy |
| Reason for referral | Pathogenic variant in ANKRD11 gene associated with KBG syndrome |

| Birth History and Vitals | |
|---|---|
| Pregnancy: | Normal prenatal care; nuchal scan and US looked normal per mother; small for gestational age |
| Birth (NVD or C-section): | Caesarian section, resided in NICU |
| Gestational Age | 37 weeks, 3 weeks early |

| | |
|---|---|
| Feeding: | Neonatal feeding difficulties, nasogastric tube for 5 days, resided in the NICU |
| Postnatal Period: | Failure to thrive; no hypoxia; floppy baby |

| *Facial abnormalities:* | |
|---|---|
| Ear: | None |
| Eye: | Orbital hypertelorism, long eyelashes, minor synophrys |
| Nose: | Upturned nose, broad nasal bridge, anteverted nares, |
| Mouth: | Macrodontia |
| Other: | Short neck, |

| | |
|---|---|
| Developmental Behavior | Mild developmental delays. Agitation. Able to write name on paper, able to write time, able to draw clock with time with difficulty; no aggressive behavior, no clear indication of autism; rigid and likes routine, gives single word responses at times; fully toilet trained, able to eat and cook for herself, good self-care but lacks skills for independent living; persistent learning and language difficulties |
| Motor Delay | Delayed motor development with walking; poor coordination |
| Speech Delay: | Delayed speech development, some slurred speech currently |
| IQ | unknown |
| Intellectual Disability: | intellectual disability, mild |
| Vision: | Ocular immaturity diagnosed at 6 months; optic nerves functional |
| Hearing: | None |
| Neurology: | 4-5 generalized tonic clonic seizures starting at 4 years old for 2 years then stopped (grand mal, made her fall and break collarbone); 2 grand mal seizures last year, headaches, hypotonia, |
| Seizures/onset: | Age 4 years, 4-5 generalized tonic clonic<br>Age 15 years, 2 grand mal seizures |
| Neuroimaging: | Neuro MRI done at 4 years and last year; MRI brain shows lateral right temporal bone lesion on squamosal petrosal suture; 2020, abnormal EEG |
| Cardiovascular: | None |
| Gastrointestinal: | Vomiting associated with headaches/possible migraines around puberty- no auras; possible cyclic vomiting; no abdominal pain |
| Respiratory: | None |
| Urogenital | None |
| Musculoskeletal: | Short stature, mild clinodactyly, hypotonia, short fingers and smaller $5^{th}$ digit, short neck, no obvious sacral dimple; no scoliosis |
| Skin/Hair/Nails: | None |
| Endocrine: | None |
| Immunological: | None |
| Allergies: | No known allergies |
| Metabolic: | None |

| | |
|---|---|
| Neoplasm: | None |
| Inheritance: | Autosomal dominant |
| Family History of congenital disorders: | None |
| Family History of intellectual disabilities | None |
| Consanguinity: | None |
| Medication History: | Lamotrigine 50mg in AM, 75mg in PM, Vitamin D |
| Services | Special education; speech and language therapy; occupational therapy |
| Family History | Older twin sisters; ages 19 years with no developmental issues, not tested for KBG syndrome |

Standardized Testing:

2010: EEG shows epileptiform discharges maximal over the frontal regions but increased in sleep
2013: EEG shows normal sleep architecture with occasional epileptiform discharges
Jan-7-2021: MRI Head shows lateral right temporal bone lesion on squamosal petrosal suture

Nov 2020- WIAT111

| WIAT III – Assessment Scores | | | | |
|---|---|---|---|---|
| Area assessed | Previous result/score | Date | Most recent result/score | Date |
| Early Reading Skills Standard Score | | | | |
| Comprehension Standard Score | 65 | 15/05/19 | 56 | 12/11/20 |
| Word Reading Standard Score | 71 | 15/05/19 | 62 | 12/11/20 |
| Oral Fluency Standard Score | | 15/05/19 | Biennial subtest | 12/11/20 |
| Oral Accuracy Standard Score | | 15/05/19 | Biennial subtest | 12/11/20 |
| Reading Speed Standard Score | Disapplied | 15/05/19 | Biennial subtest | 12/11/20 |
| Spelling Standard Score | 73 | 15/05/19 | 70 | 12/11/20 |

*Nov-2020: Clinical Evaluation of Language Fundamentals Fifth UK Edition*

| CELF 5 UK Language tests | Scaled Scores | Percentile Rank |
|---|---|---|
| Word Classes | 4 | 2 |
| Following Directions | 4 | 2 |
| Formulated Sentences | 1 | 0.1 |
| Recalling Sentences | 7 | 16 |
| Understanding Spoken Paragraphs | 3 | 1 |
| Word Definitions | 4 | 2 |
| Sentence Assembly | 2 | 1 |
| Semantic Relationships | 4 | 2 |

| | Standard Scores | Percentile Rank |
|---|---|---|
| Core Language Score | 62 | 1 |
| Receptive Language Index | 63 | 1 |
| Expressive Language Index | 61 | 0.5 |
| Language Content Index | 57 | 0.2 |
| Language Memory Index | 66 | 1 |

# Y

| | |
|---|---|
| Gene Name: | ANKRD11 |
| NM_ | NM_013277.5 |
| cDNA change: | c.3224_3227delAAAG |
| Protein change: | p.Glu1075Glyfs*242 |
| Genomic position: | g.Ch16:89349722 |
| Hg build: | Hg38 |
| Mode of inheritance: | unknown |
| Karyotype: | unknown |
| Zygosity: | Heterozygous |
| Classification: | unknown |
| Reason for referral | The proband is a 16-year-old-male, born at 32-week gestation, now with mild global developmental delay, mild intellectual disability, and multiple anomalies including pineal cyst, dental and palatal anomalies, profound unilateral hearing loss, with history of bilateral hamstring contractures, and bilateral ankle contractures, history of cryptorchidism, and history of seizures. |

| Birth History and Vitals | |
|---|---|
| Pregnancy: | Maternal diabetes; difficult pregnancy; lost 24lb during pregnancy (130 to106 lb by birth) |
| Birth (NVD or C-section): | Caesarian section |
| Gestational Age | 32 weeks; premature |
| Feeding: | Feeding difficulties in infancy |
| Postnatal Period: | Neonatal jaundice; failure to thrive; resided NICU for 8 weeks; neonatal respiratory distress; immature lungs; failed neonatal hearing test; absent eyelashes. |

| | |
|---|---|
| Birth weight (percentile): | <1% |
| Birth length: | <1% |
| Birth head circumference: | unknown |
| Current weight: | >85% 254 lbs.(overweight-attributed to medications) |
| Current length: | >85% 6' tall; |
| Current head circumference: | unknown |

| | |
|---|---|
| Developmental Behavior | ADHD; autism; anger issues; hyperactive; depression; aggression; (now on therapy); attempted suicide twice by jumping out of car; nail biting; mild learning disabled; delay in reading-technology is very helpful; has a sense of humor; mood swings; IQ every two years, ranging from 98 to 68; mother reported his IQ is 73 reads at a 2nd grade reading level. Struggled in Junior high; Emotionally and mentally at around age 10; can cook and do chores |
| Motor Delay | Gait disturbance; Botox for tight Achilles tendon; poor fine motor coordination of hands |
| Speech Delay: | No, very verbal |
| IQ | IQ73 |
| Intellectual Disability: | mild |
| Neurology: | ADHD; migraines; seizures; cerebral palsy; no hypotonia as infant; depression; autism diagnosis given as per mother. |
| Neuroimaging: | (2015); Pineal cyst; |
| *Facial abnormalities:* | Mild dysmorphia |
| Ear: | Bilateral hearing loss (wears hearing aids) |
| Eye: | Synopsys; (unibrow); absent eyelashes (as neonate); full eyebrows |
| Nose: | Anteverted nares |
| Mouth: | Persistent primary teeth; wide-spaced teeth; frenulum (tongue-tied); thin upper lip vermillion; micrognathia; retained primary teeth; overgrown gums; macrodontia; tonsils would obstruct airways in sleep |
| Other: | Short chin; strong jaw musculature |
| Vision: | Eyeglasses prescribed, not compliant in wearing them; |
| Hearing: | Hearing loss Bilateral-uses hearing aids; |
| Cardiovascular: | patent foramen ovale |
| Gastrointestinal: | Constipation; hospitalized 3-4 times for impaction; gastro immobility; GERD; episodic vomiting; dumping syndrome; neonatal jaundice; feeding difficulties in infancy; never NG tube; aspiration risk as child; |
| Respiratory: | Recurrent upper respiratory infections; asthma; reactive airway disease |
| Urogenital | Undescended testes due to inguinal hernia, (surgically resolved); cryptorchidism; renal hypertension; |
| Musculoskeletal: | Mild scoliosis, (<40 °); issues with hips, (hip dysplasia); muscles cramps and pain, never hormone treatment; gait disturbance; pes planus; toe walker; club foot treated with Botox; no tethered spinal cord |
| Skin/Hair/Nails: | Acne; dry skin; |
| Endocrine: | no |
| Immunological: | Allergies |

| | |
|---|---|
| Allergies: | penicillin; amoxicillin; clarithromycin; mold extract; lactose intolerant as infant, (now resolved) |
| Metabolic: | Inguinal hernia: sleep disorder treated with melatonin; failure to thrive in infancy; fatty liver; renal hypertension; neonatal respiratory distress, obese (due to medications); lean as a child. |
| Neoplasm: | none |
| Inheritance: | Unknown; other tested and does not carry ANKRD11 gene variant; biological father unavailable for testing; |
| Family History of congenital disorders: | Miscarriages: Father also demonstrated similar behavioral issues, per the mother. |
| Family History of intellectual disabilities | Cousins with Down syndrome |
| Consanguinity: | no |
| Medications: | Depakote, 1750 mg daily; Miralax; magnesium; Seroquel; Sertraline, melatonin at night, guanfacine, vitamin E, fish oil, Lisinopril for blood pressure. |
| Services | Early intervention since infancy; OT; PT; life skill classes; school for the deaf; received speech therapy in the past |
| Family History | Lives with mother and stepfather; has older biological half-brother with no developmental or intellectual issues; |

| | |
|---|---|
| Survey & Parental replies to determine Eosinophil Esophagitis status: | |
| Question: | Answer provided by parent: |
| 1)Any allergy diagnosed? – to what? | 1) penicillin; amoxicillin; clarithromycin; mold extract; lactose intolerant as infant, (now resolved) |
| 2)Any acute skin reaction or swelling reaction to a food? | 2)no |
| 3)Anaphylaxis | 3) no |
| 4)Any episodes of diarrhea/vomiting or failure to thrive in infancy responsive to food exclusion | 4) yes, episodic vomiting |
| 5)Was there any failure to gain weight? | 5) failure to thrive in infancy; |
| 6)Eczema – severity / extent | 6) dry skin on elbows |
| 7) Asthma – severity / preventer | 7)yes, mild asthma |
| 8) Allergic rhinitis | 8) no |
| 9) Odynophagia | 9) no |
| 10)Dysphagia; particularly meat and bread getting stuck. | 10)yes, aspiration risk as child; |
| 11) Is there a need to have food cut up very small? | 11)no, no feeding tube |

Standardized Testing: none performed for this study

# Z

| | |
|---|---|
| Gene Name: | ANKRD11 |
| NM_ | NM_013275.5 |
| cDNA change: | c.7607G>A |
| Protein change: | p.Arg2536Gln |
| Genomic position: | g. |
| Hg build: | GRch37 |
| Mode of inheritance: | Maternally inherited |
| Zygosity: | Heterozygous |
| Classification: | Likely pathogenic |
| Other variants: | |
| Reason for referral | |

This is the family discovered via ClinVar, and this is a 14-year-old female of Greek origin (living in Germany) with suspected syndromic disease, mild intellectual disability, microcephaly, familial short stature and some dysmorphic features (thick eyebrows, synophrys, prominent nasal tip, macrodontia, thin upper lip vermillion). In this case, the variant was inherited from the mother who has short stature and mild learning disabilities. The maternal grandmother has short stature and a learning disability but has not been tested for the variant. This family was not available for a videoconference, and there is thus substantial missing information. The severity of intellectual disability was never quantified via cognitive testing, but is likely in the mild range. She is noted to sometimes have abnormal mood, and she did has aggressive violent behavior as a child. This included some impulsivity, but did not include self-injurious behavior or compulsive behaviors. Her cardiac status is unknown, in terms of any possible congenital heart defects. She had delayed walking, around 17 months of age. It is not known if she had hypotonia.  She does have pes planus. She was not noted to have feeding difficulties in infancy, but she did have vomiting.
